# Durability of the BNT162b2 XBB.1.5-adapted vaccine against JN.1 hospitalisation in Europe, October 2023 to August 2024: A test-negative case-control study using the id.DRIVE platform

**DOI:** 10.1101/2025.05.02.25326904

**Authors:** HR Volkman, L de Munter, JL Nguyen, TMP Tran, M Mitratza, C Marques, L Choi, S Valluri, J Yang, A Antón, I Casas, E Conde-Sousa, L Drikite, B Grüner, G Icardi, G Luit ten Kate, C Martin, A Mira-Iglesias, A Orrico-Sánchez, S Otero-Romero, G Rohde, L Jodar, JM McLaughlin, K Bollaerts

## Abstract

**Background:** While multiple studies have shown that the BNT162b2 XBB.1.5-adapted mRNA COVID-19 vaccine (BNT162b2 XBB vaccine) is effective in preventing COVID-19 hospitalisation and death, effectiveness beyond six months remains unexplored.

**Aim:** We extended our previous study of BNT162b2 XBB vaccine effectiveness (VE) to evaluate durability against JN.1-related hospitalisation up to 46 weeks since dose using the id.DRIVE platform across Belgium, Germany, Italy, and Spain.

**Methods:** This multi-country, multi-centre test-negative case-control study assessed the effectiveness and durability of the BNT162b2 XBB vaccine in preventing JN.1-associated hospitalisation among adults with severe acute respiratory infection between October 2023 and August 2024. Each case was matched with up to four controls by symptom onset date and study site. Multivariable analyses were adjusted for symptom onset date, age, sex, number of chronic conditions, and influenza vaccination receipt.

**Results:** Among 827 test-positive cases and 2232 test-negative controls, protection against hospitalisation was sustained from 2 to <30 weeks since dose, with evidence of significant waning thereafter. VE was 64.5% (95% CI: 56.6; 71.0) at 2 to <30 weeks, and 4.9% (95% CI: −30.3; 30.7) at 30 to <46 weeks.

**Conclusions:** Despite the vaccine target not matching the predominant subvariant, BNT162b2 XBB vaccine protected against JN.1-related hospitalization for up to 30 weeks, but protection against hospitalisation was non-significant after 30 weeks since dose, potentially due to further shift in circulating SARS-CoV-2 strains and/or waning immunity. Given the high COVID-19 activity in Europe during summer 2024, an additional vaccination after six months is warranted for those at risk of COVID-19 hospitalisation to maintain year-round protection.

## Introduction

Since its authorisation by the European Medicines Agency (EMA) on 30 August 2023 [1], numerous real-world studies have reported on the vaccine effectiveness (VE) of the BNT162b2 XBB.1.5-adapted mRNA COVID-19 vaccine (Pfizer/BioNTech 2023–2024 formulation, hereafter referred to as the BNT162b2 XBB vaccine) during the 2023–2024 season [2–20]. These studies collectively show that the BNT162b2 XBB vaccine is effective against severe endpoints such as hospitalisation [2–5, 8, 9, 11, 12, 15–18], ICU admission [4], and death [3, 17]. However, its effectiveness in preventing hospitalisation is likely lower against Omicron subvariant JN.1 than XBB (VE of 61%-76% during XBB predominance and VE of 35%-54% during JN.1 predominance) [2–5, 9, 11, 12, 17].

Despite this broad evidence base, the long-term durability of XBB-adapted COVID-19 vaccines beyond 6 months since dose remains unknown. This gap in evidence on long-term effectiveness is especially relevant considering that from June–August 2024, many countries in Europe and around the globe experienced an increase in COVID-19 hospitalisations [21], long after Northern Hemisphere autumn/winter-focused COVID-19 vaccination campaigns had concluded. Changes in COVID-19 dynamics, including SARS-CoV-2 strain shifts [22], complex and evolving population immunity [23], and aseasonal/acyclical increases in hospitalisations [24], have continued since the conclusion of the World Health Organization (WHO)-declared global health emergency [25]. Therefore, it is critical for public health decision-makers to have timely information on the long-term durability of COVID-19 vaccines.

Therefore, we sought to extend the study period of our previously published real-world effectiveness analysis of the BNT162b2 XBB vaccine against JN.1-associated hospitalisation across four European countries (Belgium, Germany, Italy, and Spain) [12]. Our previous report indicated that the BNT162b2 XBB vaccine was 54.8% (95% CI: 39.7; 66.0) effective against JN.1-related hospitalisation after a median of 63 days following vaccination, with no evidence of waning protection through five months [12]. In this analysis, we measured the long-term durability of the BNT162b2 XBB vaccine, both overall and in risk groups, up to 46 weeks post-dose using the id.DRIVE study platform during a period of JN.1 predominance.

## Methods

### Study design

As previously described in our analysis of BNT162b2 XBB VE through April 2024 [12], id.DRIVE is an ongoing multi-country, multi-centre test-negative case-control study that began in Europe in November 2020. The study aims to estimate VE against COVID-19-related hospitalisations with laboratory confirmation of SARS-CoV-2 infection among patients admitted with severe acute respiratory infections (SARI). The id.DRIVE Master protocol (EUPAS42328) was approved by independent ethics committees (IECs) at each study site. Patients with SARI were enrolled both prospectively and retrospectively across eight sites, including six hospitals and two hospital networks across Belgium, Germany, Italy, and Spain from 2 October 2023 to 31 August 2024 (**Supplementary Table S1**).

### Study objective

The primary objective was to determine the VE and long-term durability of the BNT162b2 XBB vaccine in preventing COVID-19 hospitalisations through 31 August 2024 (up to 46 weeks since dose).

### Study participants

Patients included adults aged ≥18 years eligible for COVID-19 vaccination based on national or regional guidelines who were hospitalised for ≥1 night and diagnosed with SARI, based on the European Centre for Disease Prevention and Control (ECDC) case definition [26]. Qualifying SARI symptoms included cough, objective fever (≥38⁰C), shortness of breath, anosmia, ageusia, or dysgeusia. Symptom onset date must have occurred ≤14 days before hospital admission. Patients were excluded if admitted to the hospital for COVID-19 within the past 3 months, were incapable of providing a nasopharyngeal or oropharyngeal specimen, or had received a COVID-19 vaccine dose from a manufacturer not authorised by the EMA. Informed consent was obtained from patients or their legal representatives except at study sites where IECs authorised waivers of consent.

### SARS-CoV-2 laboratory testing and variant identification

All patients were tested for SARS-CoV-2 by reverse transcription polymerase chain reaction (RT-PCR) or multiplex PCR between 14 days prior to, and up to 72 hours following, hospital admission. The SARS-CoV-2 infecting strain was further characterised in positive specimens using whole genome sequencing or next-generation sequencing. When sequencing was not possible due to insufficient viral load or poor sample quality, the predominant subvariant present in the country of hospitalisation on symptom onset date was imputed from national reporting sources.

A predominance threshold of ≥70% of sequenced samples was established using country-level genomic reporting by calendar time. Sub-lineages of XBB or JN.1 (such as EG.5, KP.2, and KP.3) were collapsed into their parent lineages. When neither XBB nor JN.1 reached the predominance threshold of ≥70%, the time was classified as a transition period. Country-specific XBB and JN.1 predominance periods are defined in **Supplementary Table S1**.

### Exposure and outcome definition

Exposed patients received at least one dose of the BNT162b2 XBB vaccine, irrespective of prior COVID-19 vaccination status, at least 14 days before symptom onset and at least 12 weeks after receipt of any prior COVID-19 vaccine. The unexposed group included patients who did not receive any COVID-19 vaccine during the 2023–2024 season, including never vaccinated patients and those who received COVID-19 vaccines prior to the 2023–2024 season and at least 12 weeks before symptom onset, following national eligibility guidelines [27].

’Test-positive cases’ were patients who met the SARI case definition and had at least one positive SARS-CoV-2 result by PCR. ‘Test-negative controls’ were patients who also met the SARI case definition and received a negative result for all SARS-CoV-2 tests performed.

### Data collection and management

COVID-19 vaccination data were collected from registries, medical records, or vaccination cards. Data were collected on age, sex, predefined chronic conditions associated with an increased risk of severe outcomes from SARS-CoV-2 infection (**Supplementary Table S2**), body mass index (BMI), symptom onset date, and clinical course of the hospitalisation. Trained study staff entered data from medical records and patient interviews into an electronic Case Report Form (eCRF, Castor^®^). Pseudonymised data were transferred to a secure central research server hosted by P95 Clinical and Epidemiology Services (Leuven, Belgium) for analysis.

### Statistical methods

Odds ratios (OR) comparing the odds of vaccination between test-positive cases and test-negative controls were estimated using multivariable Generalized Estimating Equation (GEE) logistic regression models. These models accounted for potential intra-cluster correlation among patients from the same study site [28]. The variances associated with ORs were calculated using a robust sandwich estimator, yielding a consistent and asymptotically normal estimator [29]. VE was calculated as (1–OR) × 100%. Models adjusted for symptom onset date, age, sex, number of chronic conditions, and receipt of influenza vaccination in the past 12 months. Each case was matched with up to four controls by study site and 14-day interval from the earliest onset date to minimise potential residual time-varying confounding. Matching was conducted independently for each study objective, using distinct datasets. Calendar time for symptom onset date was modelled using a cubic spline, with the number of knots aligned with the number of symptom onset months included in the analysis. The effect of age was captured using a cubic spline with two predefined knots at ages 50 and 65 years. When <10% of patients in the dataset were <50 years of age, a single knot at age 65 was used. The cutoff points used to define the age group categories determined the selection of knots for the age effect. To overcome multicollinearity while fitting GEE models, we applied penalisation for spline terms of symptom onset date utilising a modified Penalized GEE (PGEE) package with a ridge penalty term [30].

To estimate VE by time since the last BNT162b2 XBB vaccine dose, the logistic regression model was extended to include the interaction between exposure status and time since the last vaccine dose. The effect of time since the last vaccine dose was captured by a cubic regression spline with a knot at 182 days (or 26 weeks), based on prior evidence of waning VE after 26 weeks for any XBB vaccine [31]. The time-dependent OR was derived on a one-dimensional grid ranging from the minimum to the maximum values of time since the last dose, with a step size of 1 day. The sandwich estimator was used to calculate the variance of the VE by time since the last vaccine dose.

Several sensitivity analyses were performed to assess the robustness of the results obtained from the PGEE approach. These analyses considered different choices regarding the number and location of knots for both symptom onset date and age, the impact of prior COVID-19 infection, and the impact of removing adjustment for influenza vaccination status.

Only patients with complete data on exposure status, SARS-CoV-2 test results, and relevant confounders were included in regression analyses [32]. As 1.6% (113/7223) of the patients with SARI were excluded from analysis due to missing chronic conditions or missing influenza vaccination history (**Figure 2**), the complete case analysis is considered unlikely to produce biased estimates.

Sample size calculations, model specifications, and fitting procedures are provided in the supplementary materials (**Appendix A.1, A.2,** and **A.3**). All analyses were conducted using R version 4.1.2 [33].

## Results

### Study population

From 2 October 2023 to 31 August 2024, 7223 patients hospitalised with SARI were enrolled. There were 801 patients with symptom onset during XBB predominance or the transition period, and/or with non-JN.1 sequencing results who were excluded. Overall, 6094 (84.4%) patients with a confirmed JN.1 sequencing result or a SARI onset date during the country-specific JN.1 predominance period were eligible for analysis. After matching, a total of 3059 patients were ultimately included (**Supplementary Figure S1**).

Figure 1 indicates an increase in enrolment among patients with SARI overall and among test-positive cases in the winter months, which peaked in late December 2023. A subsequent increase among only test-positive cases peaked again in July 2024, corresponding to the summer increase of COVID-19 observed in Europe [34]. During JN.1 predominance, 77.4% (2368/3059) of all hospitalised patients with SARI were ≥65 years of age, 53.3% (1630/3059) were male, and 41.4% (1266/3059) had ≥3 chronic conditions (**Table 1**). The most prevalent chronic conditions included hypertension (62.4% of total patients), cardiovascular disease (41.9%), lung disease (35.6%), type 2 diabetes (30.4%), and immunodeficiency or cancer (21.2%). The median age of test-positive cases was 78 years (interquartile range [IQR] 66–86), and more than half were male (53.2%, n=440), similar to test-negative controls.

**Figure 1.**
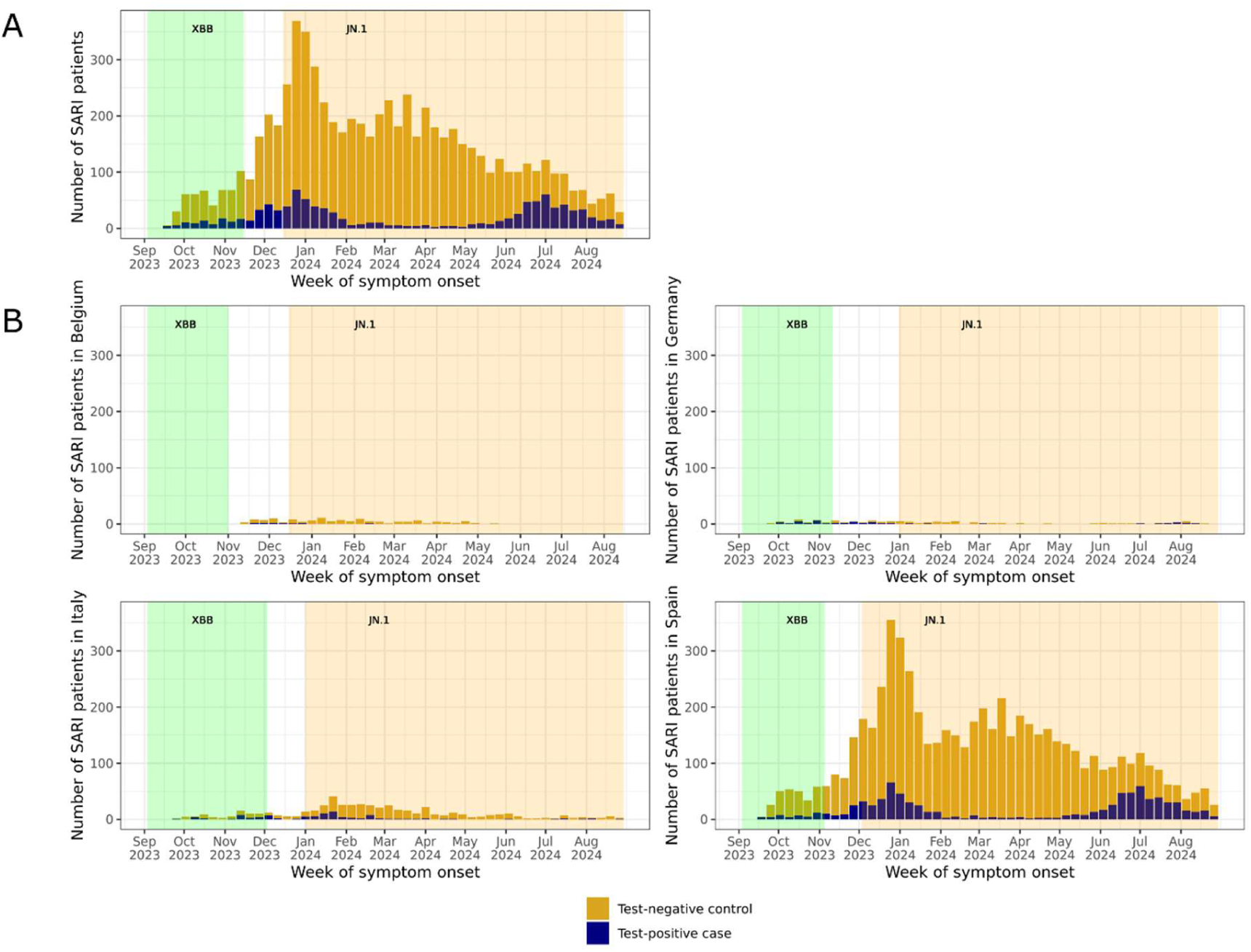
Number of enrolled patients with SARI according to SARS-CoV-2 case classification over time during XBB predominance, a transition period, and JN.1 predominance: A) overall and B) by country

**Figure 2.**
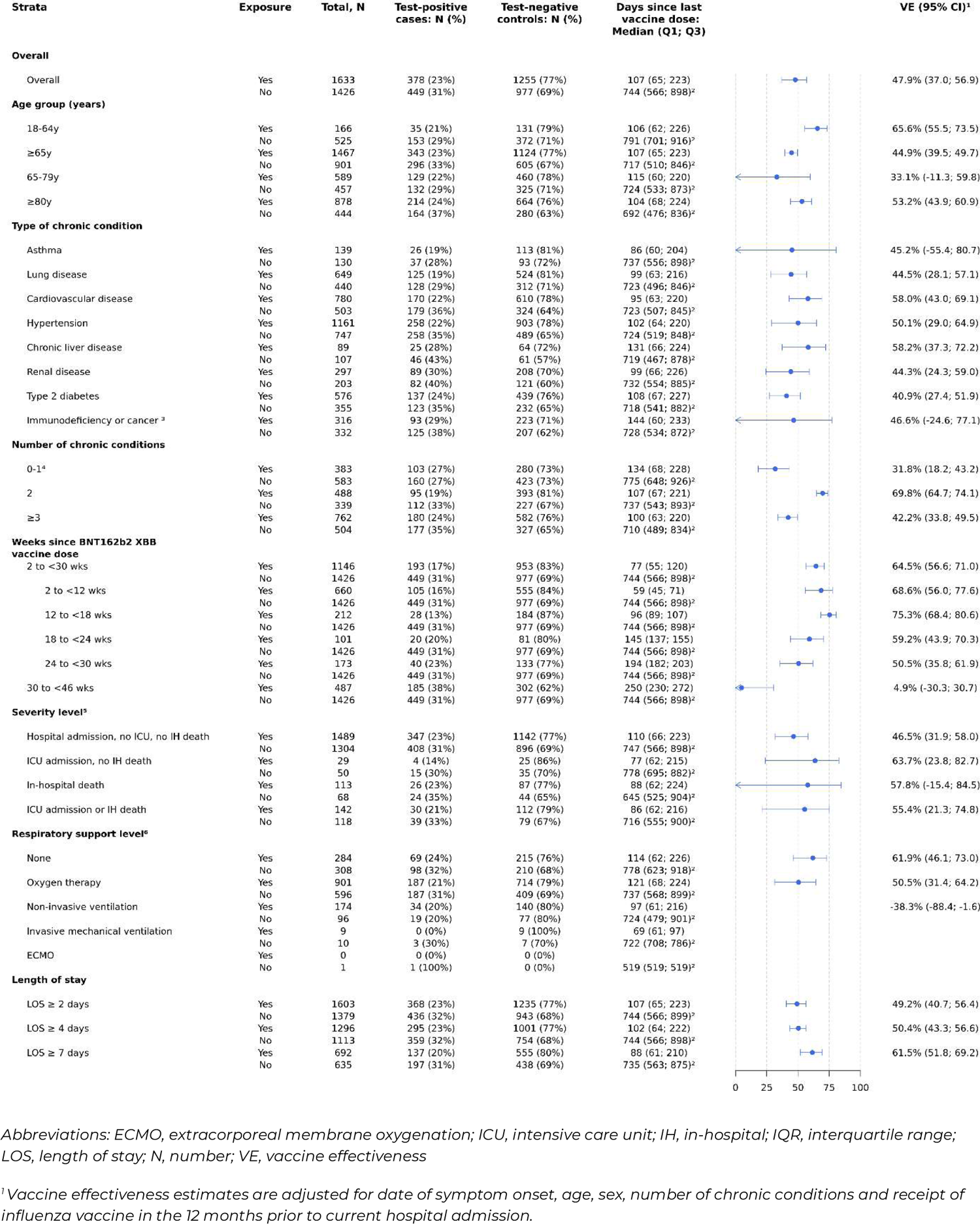
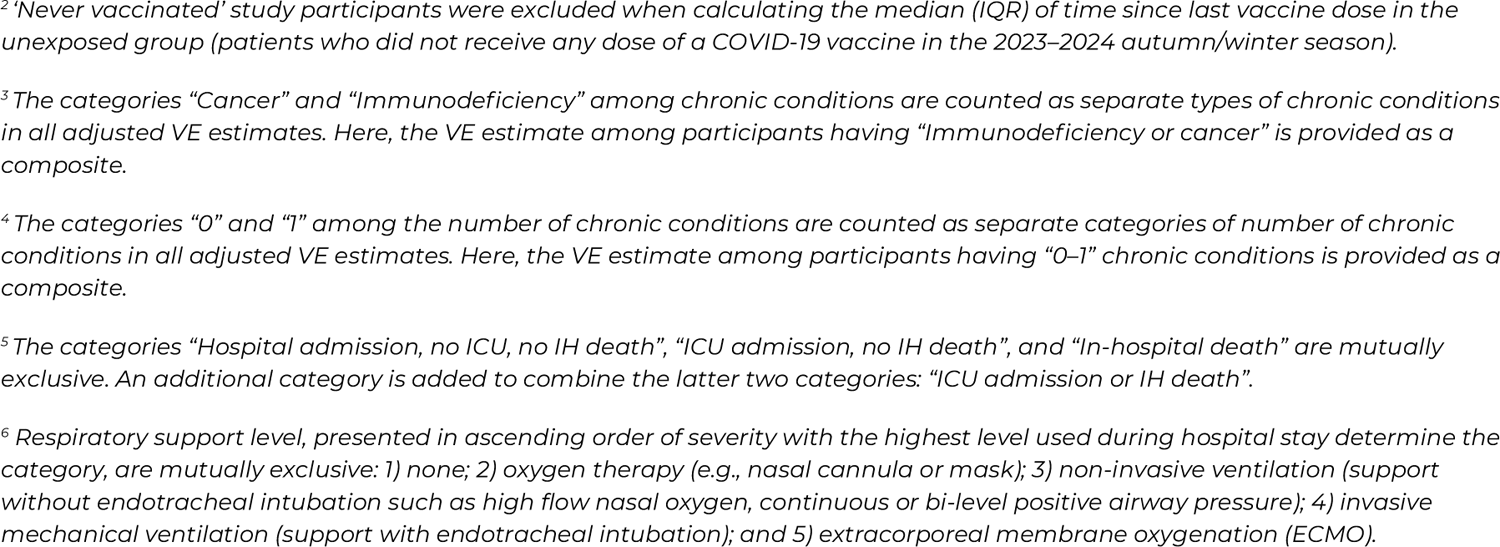
Vaccine effectiveness against JN.1-related hospitalisation in SARI patients who received at least one dose of BNT162b2 XBB vaccine compared to patients who did not receive any dose of a COVID-19 vaccine in the 2023–2024 autumn/winter season

**Table 1.** Characteristics of study participants according to receipt of BNT162b2 XBB vaccine and SARS-CoV-2 case classification.

|  | Total, N (col %) | Received BNT162b2 XBB vaccine in 2023–2024 season, (N, col %) |  | Did not receive any COVID-19 vaccine in 2023–2024 season, (N, col %) |  |
| --- | --- | --- | --- | --- | --- |
|  |  | Test-positive cases | Test-negative controls | Test-positive cases | Test-negative controls |
| **Total** | 3059 | 378 | 1255 | 449 | 977 |
| **Prior COVID-19 vaccination** |  |  |  |  |  |
| Never vaccinated | 170 (5.6) | 2 (0.5) | 7 (0.6) | 45 (10.0) | 116 (11.9) |
|  |  | Test-positive cases | Test-negative controls | Test-positive cases | Test-negative controls |
| ≥2 mRNA wildtype doses only | 914 (29.9) | 54 (14.3) | 163 (13.0) | 219 (48.8) | 478 (48.9) |
| ≥1 mRNA original bivalent BA.4/5 booster | 1732 (56.6) | 311 (82.3) | 1042 (83.0) | 134 (29.8) | 245 (25.1) |
| Other<sup>1</sup> | 243 (7.9) | 11 (2.9) | 43 (3.4) | 51 (11.4) | 138 (14.1) |
| **Sex** |  |  |  |  |  |
| Female | 1429 (46.7) | 165 (43.7) | 594 (47.3) | 222 (49.4) | 448 (45.9) |
| Male | 1630 (53.3) | 213 (56.3) | 661 (52.7) | 227 (50.6) | 529 (54.1) |
| **Age, years** |  |  |  |  |  |
| Median (IQR) | 77.0 (66.0, 86.0) | 81.0 (73.0, 88.0) | 80.0 (72.5, 88.0) | 74.0 (60.0, 84.0) | 70.0 (56.0, 80.0) |
| 18–49 | 261 (8.5) | 3 (0.8) | 19 (1.5) | 60 (13.4) | 179 (18.3) |
| 50–64 | 430 (14.1) | 32 (8.5) | 112 (8.9) | 93 (20.7) | 193 (19.8) |
| ≥65 | 2368 (77.4) | 343 (90.7) | 1124 (89.6) | 296 (65.9) | 605 (61.9) |
| **Body mass index (BMI)<sup>2</sup>, kg/m<sup>2</sup>** |  |  |  |  |  |
| Median (IQR) | 25.7 (22.8, 29.4) | 25.8 (22.8, 29.1) | 26.1 (23.0, 30.1) | 25.0 (22.6, 29.0) | 25.4 (22.7, 29.3) |
| Underweight (BMI <18.5) | 98 (4.2) | 15 (4.9) | 33 (3.2) | 21 (6.6) | 29 (4.2) |
| Normal weight (BMI ≥18.5 to <25.0) | 902 (38.4) | 115 (37.6) | 383 (37.3) | 130 (40.8) | 274 (39.4) |
| Overweight (BMI ≥25.0 to <30.0) | 752 (32.0) | 108 (35.3) | 332 (32.3) | 89 (27.9) | 223 (32.1) |
| Obese (BMI ≥30.0)<sup>3</sup> | 595 (25.4) | 68 (22.2) | 279 (27.2) | 79 (24.8) | 169 (24.3) |
| Missing | 712 | 72 | 228 | 130 | 282 |
| **Number of chronic conditions** |  |  |  |  |  |
| 0–1<sup>4</sup> | 966 (31.6) | 103 (27.2) | 280 (22.3) | 160 (35.6) | 423 (43.3) |
| 2 | 827 (27.0) | 95 (25.1) | 393 (31.3) | 112 (24.9) | 227 (23.2) |
| ≥3 | 1266 (41.4) | 180 (47.6) | 582 (46.4) | 177 (39.4) | 327 (33.5) |
| **Type of chronic condition** |  |  |  |  |  |
| Asthma | 269 (8.8) | 26 (6.9) | 113 (9.0) | 37 (8.2) | 93 (9.5) |
| Lung disease | 1089 (35.6) | 125 (33.1) | 524 (41.8) | 128 (28.5) | 312 (31.9) |
| Cardiovascular disease | 1283 (41.9) | 170 (45.0) | 610 (48.6) | 179 (39.9) | 324 (33.2) |
| Hypertension | 1908 (62.4) | 258 (68.3) | 903 (72.0) | 258 (57.5) | 489 (50.1) |
| Renal disease | 196 (6.4) | 25 (6.6) | 64 (5.1) | 46 (10.2) | 61 (6.2) |
| Liver disease | 500 (16.3) | 89 (23.5) | 208 (16.6) | 82 (18.3) | 121 (12.4) |
| Type 2 diabetes | 931 (30.4) | 137 (36.2) | 439 (35.0) | 123 (27.4) | 232 (23.7) |
|  |  | Test-positive cases | Test-negative controls | Test-positive cases | Test-negative controls |
| Immunodeficiency or cancer<sup>5</sup> | 648 (21.2) | 93 (24.6) | 223 (17.8) | 125 (27.8) | 207 (21.2) |
| **Reported prior SARS-CoV-2 infection<sup>2</sup>** |  |  |  |  |  |
| Yes | 913 (34.7) | 124 (36.3) | 396 (35.4) | 119 (31.6) | 274 (34.5) |
| No | 1717 (65.3) | 218 (63.7) | 722 (64.6) | 257 (68.4) | 520 (65.5) |
| Missing | 429 | 36 | 137 | 73 | 183 |
| **Influenza vaccination within 12 months prior to hospitalisation** |  |  |  |  |  |
| Yes | 1861 (60.8) | 358 (94.7) | 1195 (95.2) | 116 (25.8) | 192 (19.7) |
| No | 1198 (39.2) | 20 (5.3) | 60 (4.8) | 333 (74.2) | 785 (80.3) |
| **Symptom onset, month** |  |  |  |  |  |
| Nov 2023 | 4 (0.1) | 0 (0.0) | 0 (0.0) | 4 (0.9) | 0 (0.0) |
| Dec 2023 | 744 (24.3) | 59 (15.6) | 300 (23.9) | 99 (22.0) | 286 (29.3) |
| Jan 2024 | 713 (23.3) | 59 (15.6) | 334 (26.6) | 96 (21.4) | 224 (22.9) |
| Feb 2024 | 183 (6.0) | 14 (3.7) | 84 (6.7) | 24 (5.3) | 61 (6.2) |
| Mar 2024 | 121 (4.0) | 8 (2.1) | 55 (4.4) | 14 (3.1) | 44 (4.5) |
| Apr 2024 | 80 (2.6) | 10 (2.6) | 45 (3.6) | 6 (1.3) | 19 (1.9) |
| May 2024 | 206 (6.7) | 21 (5.6) | 134 (10.7) | 17 (3.8) | 34 (3.5) |
| Jun 2024 | 384 (12.6) | 71 (18.8) | 129 (10.3) | 69 (15.4) | 115 (11.8) |
| Jul 2024 | 408 (13.3) | 92 (24.3) | 108 (8.6) | 90 (20.0) | 118 (12.1) |
| Aug 2024 | 216 (7.1) | 44 (11.6) | 66 (5.3) | 30 (6.7) | 76 (7.8) |
| **Country of hospitalisation** |  |  |  |  |  |
| Belgium | 5 (0.2) | 1 (0.3) | 4 (0.3) | 0 (0.0) | 0 (0.0) |
| Germany | 25 (0.8) | 0 (0.0) | 1 (0.1) | 8 (1.8) | 16 (1.6) |
| Italy | 255 (8.3) | 15 (4.0) | 30 (2.4) | 53 (11.8) | 157 (16.1) |
| Spain | 2774 (90.7) | 362 (95.8) | 1220 (97.2) | 388 (86.4) | 804 (82.3) |
| **Length of hospital stay, days** |  |  |  |  |  |
| Median (IQR) | 6.0 (4.0, 9.0) | 5.0 (4.0, 8.0) | 6.0 (4.0, 9.0) | 6.0 (4.0, 10.0) | 6.0 (4.0, 10.0) |
| 1–3 | 650 (21.2) | 83 (22.0) | 254 (20.2) | 90 (20.0) | 223 (22.8) |
| 4–6 | 1082 (35.4) | 158 (41.8) | 446 (35.5) | 162 (36.1) | 316 (32.3) |
| 7–13 | 918 (30.0) | 99 (26.2) | 410 (32.7) | 118 (26.3) | 291 (29.8) |
| 14–27 | 320 (10.5) | 30 (7.9) | 123 (9.8) | 63 (14.0) | 104 (10.6) |
|  |  | Test-positive cases | Test-negative controls | Test-positive cases | Test-negative controls |
| 28–41 | 54 (1.8) | 7 (1.9) | 14 (1.1) | 9 (2.0) | 24 (2.5) |
| ≥42 | 35 (1.1) | 1 (0.3) | 8 (0.6) | 7 (1.6) | 19 (1.9) |
| **Level of respiratory support<sup>2</sup>** |  |  |  |  |  |
| None | 592 (24.9) | 69 (23.8) | 215 (19.9) | 98 (31.8) | 210 (29.9) |
| Oxygen therapy | 1497 (62.9) | 187 (64.5) | 714 (66.2) | 187 (60.7) | 409 (58.2) |
| Non-invasive ventilation | 270 (11.3) | 34 (11.7) | 140 (13.0) | 19 (6.2) | 77 (11.0) |
| Invasive mechanical ventilation | 19 (0.8) | 0 (0.0) | 9 (0.8) | 3 (1.0) | 7 (1.0) |
| ECMO | 1 (0.0) | 0 (0.0) | 0 (0.0) | 1 (0.3) | 0 (0.0) |
| Missing | 680 | 88 | 177 | 141 | 274 |
| **Level of SARI severity<sup>2</sup>** |  |  |  |  |  |
| Hospital admission without ICU admission and without in-hospital death | 2793 (91.5) | 347 (92.0) | 1142 (91.1) | 408 (91.3) | 896 (91.9) |
| ICU admission without in-hospital death | 79 (2.6) | 4 (1.1) | 25 (2.0) | 15 (3.4) | 35 (3.6) |
| In-hospital death | 181 (5.9) | 26 (6.9) | 87 (6.9) | 24 (5.4) | 44 (4.5) |
| Missing | 6 | 1 | 1 | 2 | 2 |

Abbreviations: ECMO, extracorporeal membrane oxygenation; ICU, intensive care unit; IQR, interquartile range; N, number.
<sup>1</sup> "Other" includes the following: mRNA original bivalent BA.1 vaccine, a single mRNA wildtype dose only, a single mRNA wildtype dose following non-mRNA dose(s), ≥2 mRNA wildtype doses following non-mRNA doses, any number of non-mRNA doses.
<sup>2</sup> Percentages exclude study participants with missing values.
<sup>3</sup> Obese classification was based on both BMI ≥30.0 kg/m<sup>2</sup> and obesity as a categorical variable (yes/no).
<sup>4</sup> The categories "0" and "1" among the number of chronic conditions are counted as separate categories of number of chronic conditions, based on the list of chronic conditions provided in Supplementary Table S2. Here, the cell counts among participants having "0–1" chronic conditions are provided as a composite.
<sup>5</sup> The categories "Cancer" and "Immunodeficiency" among chronic conditions are identified as separate types of chronic conditions. Here, the cell counts among participants having "Immunodeficiency or cancer" are provided as a composite.

In total, 53.4% (1633/3059) of participants received the BNT162b2 XBB vaccine, including 45.7% (378/827) of test-positive cases and 56.2% (1255/2232) of test-negative controls (**Table 1**). Among the 1426 patients who did not receive a COVID-19 vaccine during the 2023–2024 season, 161 (11.3%) had never been vaccinated, 697 (48.9%) received ≥2 mRNA wildtype doses only, and 379 (26.6%) most recently received an original bivalent BA.4/5-adapted booster. Patients who were not vaccinated during the 2023–2024 season tended to be younger compared with those who received the BNT162b2 XBB vaccine (median age 71 years [IQR: 57–82] versus 80 years [IQR: 73–88]) and had fewer chronic conditions (35.3% with ≥3 chronic conditions compared to 46.7%).

The median length of hospital stay was 6 days (IQR: 4–9) (**Table 1**). In total, 5.9% of patients (181/3059) died in the hospital. One-quarter (24.9% [592/2379]) of patients did not require respiratory support, including 27.9% (167/598) of test-positive cases and 23.9% (425/1781) of test-negative controls. Overall, 62.9% (1497/2379) of patients received oxygen therapy, 11.3% (270/2379) received non-invasive ventilation, and 0.8% (20/2379) received invasive mechanical ventilation or extracorporeal membrane oxygenation (ECMO).

### COVID-19 vaccine effectiveness

The overall effectiveness of the BNT162b2 XBB vaccine against JN.1-related hospitalisation was 47.9% (95% CI: 37.0; 56.9) at a median of 107 (IQR: 65–223) days since dose compared with no COVID-19 vaccination during the 2023– 2024 season (Figure 2).

Effectiveness was higher among patients 18–64 years of age than those ≥65 years (65.6% [95% CI: 55.5; 73.5] and 44.9% [95% CI: 39.5; 49.7], respectively). VE was similar when stratified across a broad range of chronic condition groups; however, when considering the number of chronic conditions, patients with 2 chronic conditions had a higher VE than patients with 0–1 or ≥3 chronic conditions. VE was higher against more severe in-hospital endpoints, such as ICU admission or death than hospital admission alone, although confidence intervals overlapped. VE against hospital admission without ICU admission or death was 46.5% (95% CI: 31.9; 58.0). VE against ICU admission or in-hospital death was 55.4% (95% CI: 21.3; 74.8). The sample size was insufficient to estimate VE for the more severe respiratory support endpoints, such as invasive mechanical ventilation or ECMO. Effectiveness appeared slightly higher against longer hospital stays, although confidence intervals overlapped, with VE against a JN.1-related hospitalisation of ≥2 days was 49.2% (95% CI: 40.7 to 56.4), while VE against a hospitalisation with a length of ≥7 days was 61.5% (95% CI: 51.8; 69.2) (Figure 2).

To determine BNT162b2 XBB VE according to prior COVID-19 vaccination status and to discern any potential residual protection from COVID-19 vaccines received before the 2023–2024 season, VE was stratified by distinct comparison groups (**Supplementary Figure S2**). Compared with receiving ≥1 mRNA BA.4/5 original bivalent dose, the BNT162b2 XBB vaccine was 48.6% (95% CI: 38.9; 56.7) effective, similar to the overall estimate that used a comparison group of no vaccine in the 2023–2024 season. When compared with ≥2 mRNA wildtype doses only, VE was also similar to the overall VE estimate at 46.4% (95% CI: 37.1 to 54.4). VE of the BNT162b2 XBB vaccine was lower when only compared to patients never vaccinated against COVID-19 at 26.7% (95% CI: −38.8; 61.3), although wide confidence intervals suggested considerable uncertainty of the point estimate.

### Durability

VE stratified by time since BNT162b2 XBB vaccine dose was 64.5% (95% CI: 56.6; 71.0) at 2 to <30 weeks, with a VE of 68.6% (95% CI: 56.0; 77.6) at 2 to <12 weeks, 75.3% (95% CI: 68.4; 80.6) at 12 to <18 weeks, 59.2% (95% CI: 43.9; 70.3) at 18 to <24 weeks, and 50.5% (95% CI: 35.8; 61.9) at 24 to <30 weeks (Figure 2). VE at 30 to <46 weeks since BNT162b2 XBB vaccine dose was 4.9% (95% CI: −30.3; 30.7). These stratified results are consistent with estimates from the spline-based approach (Figure 3), which indicated a slight increase in VE from approximately 2 to 15 weeks since receipt of the BNT162b2 XBB vaccine, followed by relatively flat VE through 25 weeks, and lastly, a steeper decrease in VE after 25 weeks that reached a central estimate of 0% effectiveness at 33 weeks since dose.

**Figure 3.**
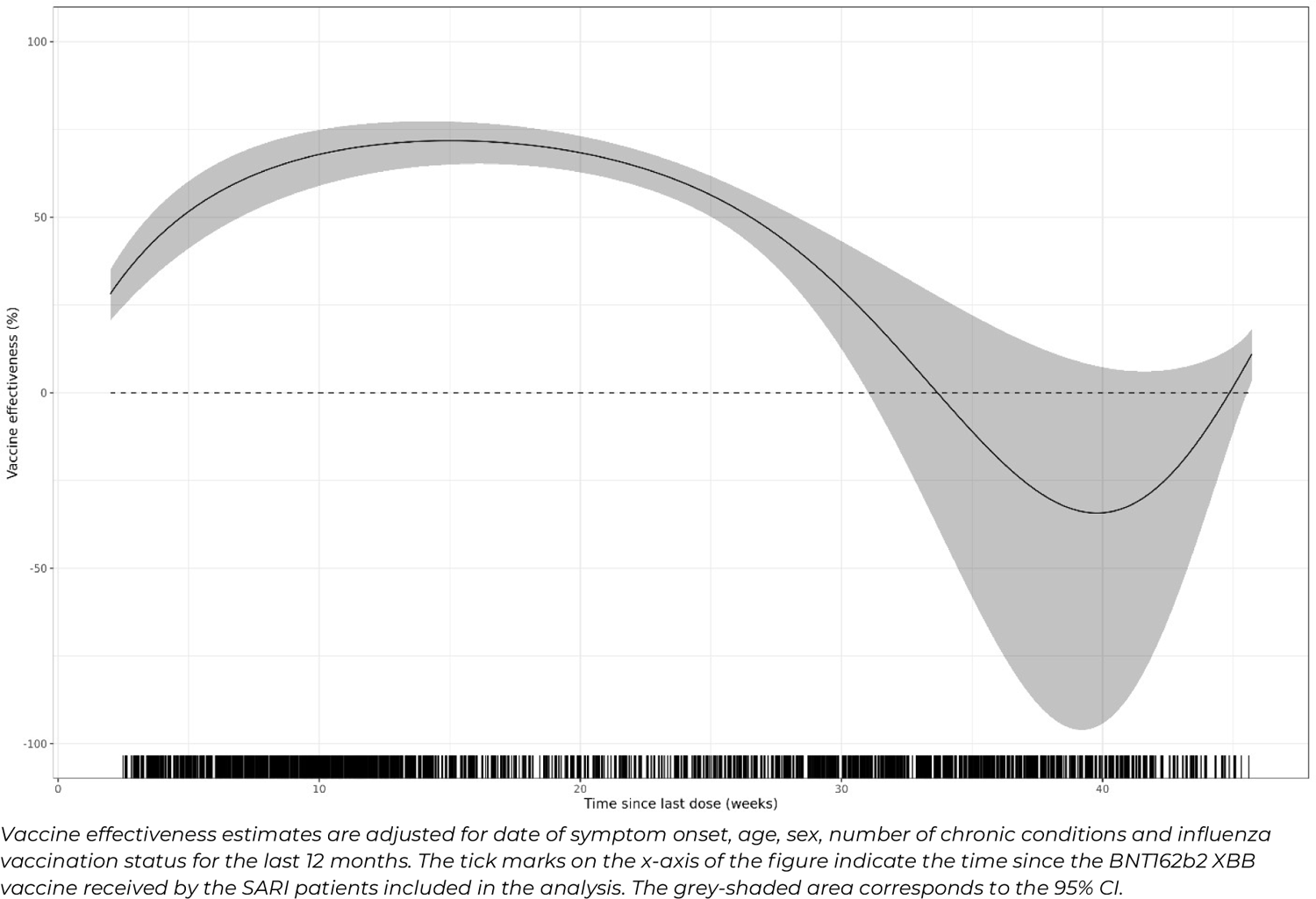
Spline-based vaccine effectiveness by time since dose against JN.1-related hospitalisation among SARI patients who received at least one dose of BNT162b2 XBB vaccine compared to patients who did not receive any dose of a COVID-19 vaccine in the 2023–2024 autumn/winter season.

When stratified by age groups and number of chronic conditions (Figure 4 and Figure 5), the durability of the BNT162b2 XBB vaccine followed a waning pattern consistent with the overall trend wherein effectiveness begins decreasing in the 18 to <24 weeks post-dose stratum. Similar results were observed when stratified by level of SARI severity (**Supplementary Figure S3**).

**Figure 4.**
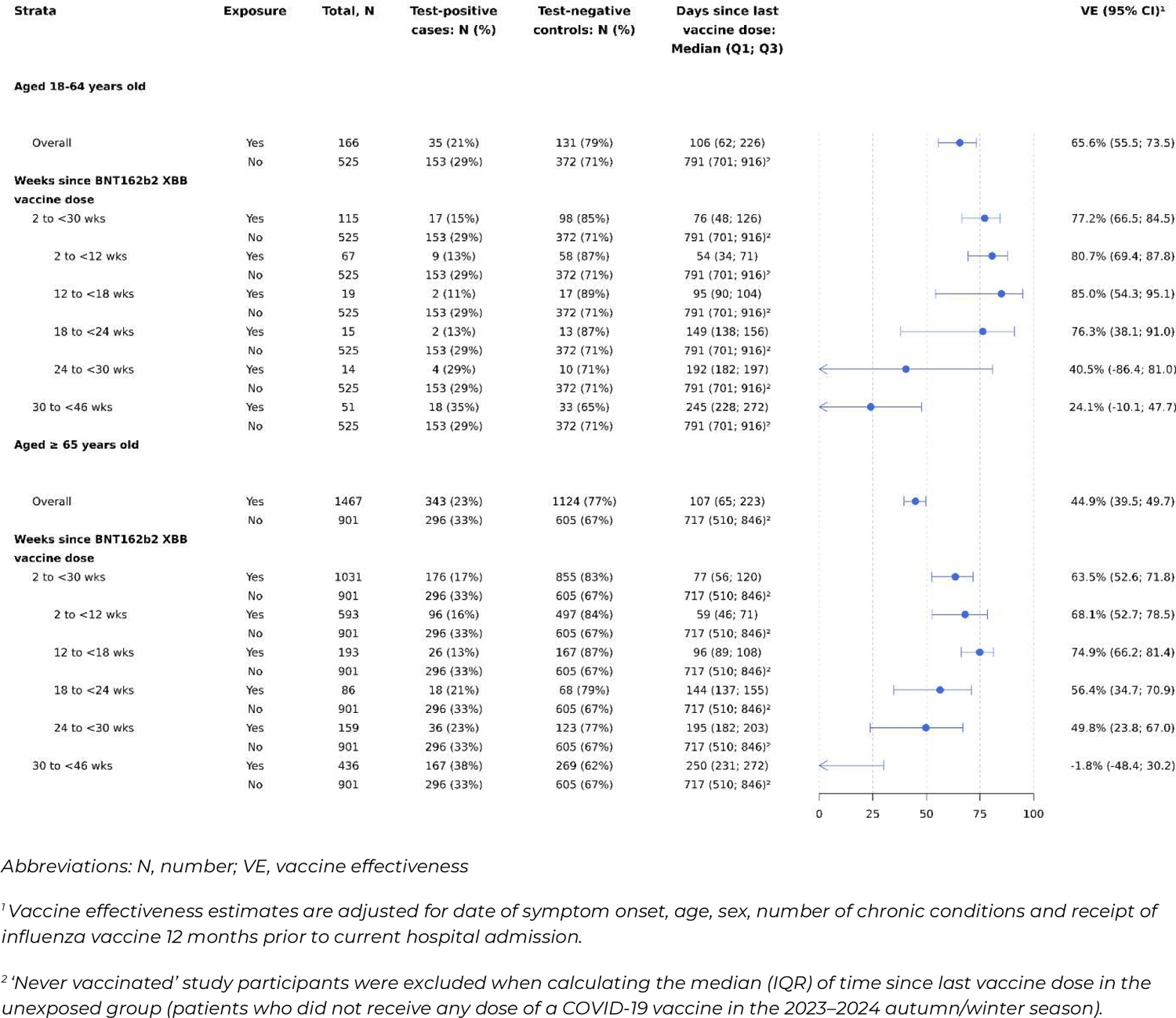
Vaccine effectiveness against JN.1-related hospitalisation in SARI patients who received at least one dose of BNT162b2 XBB vaccine compared to patients who did not receive any dose of a COVID-19 vaccine in the 2023–2024 autumn/winter, with durability stratified by age group.

**Figure 5.**
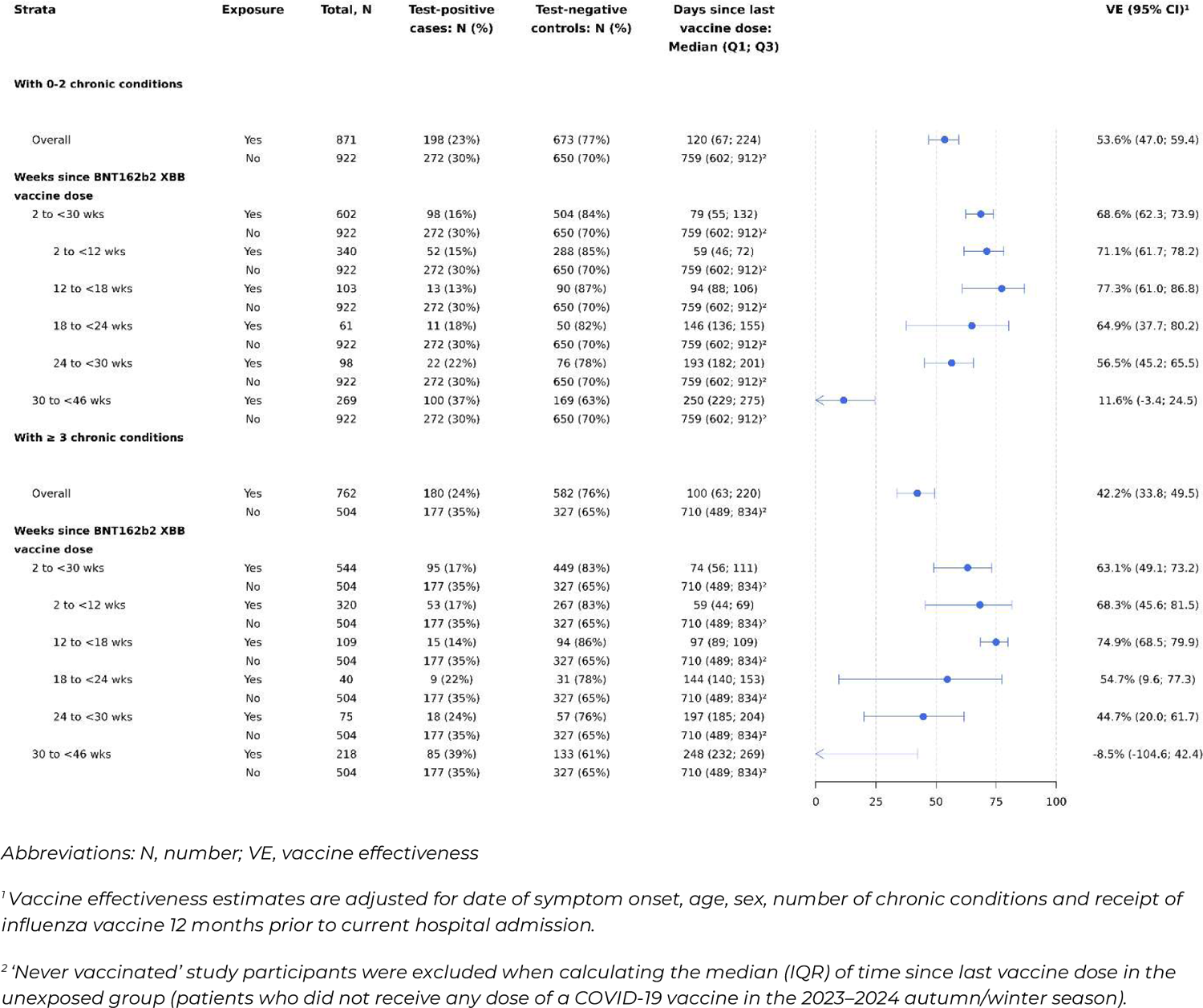
Vaccine effectiveness against JN.1-related hospitalisation in SARI patients who received at least one dose of BNT162b2 XBB vaccine compared to patients who did not receive any dose of a COVID-19 vaccine in the 2023–2024 autumn/winter, with durability stratified by number of chronic conditions.

### Sensitivity analyses

Results of sensitivity analyses that changed the number and location of knots for both symptom onset date and age were comparable to VE estimates obtained in the main analysis. When stratified by prior COVID-19 infection status, VE was 39.8% (95% CI: 12.4; 58.6) among patients with documented prior COVID-19 infection and 46.4% (95% CI: 36.0; 55.1) among patients with no documented prior COVID-19 infection (**Supplementary Figure S8**).

Estimates were lower after removing adjustment for influenza vaccination status; VE was 40.9% (95% CI: 24.3; 53.9) without influenza vaccination status adjustment (**Supplementary Figure S9**) and 47.9% (95% CI: 37.0; 56.9) with influenza vaccination status adjustment (Figure 2).

## Discussion

In this test-negative case-control study conducted in the id.DRIVE platform in Europe, the BNT162b2 XBB vaccine was 47.9% (95% CI: 37.0; 56.9) effective at a median of 107 days (IQR: 65; 223) since dose against JN.1-associated hospitalisation. VE peaked at 10–12 weeks since dose, with a decline observed after 15 weeks, and continuing to drop through 24 to <30 weeks since dose, irrespective of age group and comorbidity status. We found no evidence of protection against JN.1-associated hospitalisation beyond 30 weeks since dose.

The VE patterns observed in this study are consistent with the limited available evidence on the durability of XBB-adapted vaccines. A study conducted in Europe by Nunes and colleagues of primarily BNT162b2 XBB vaccine during JN.1 predominance, found that VE against hospitalisation was 47% (95% CI: 32; 59) for adults aged 65–79 years and 39% (95% CI: 17; 54) for adults aged ≥80 years at 90 to 179 days (∼13 to 25.5 weeks) since dose, with no evidence of waning through 25.5 weeks [17]. In our study, we observed a numerically higher VE of 56.4% (95% CI: 34.7; 70.9) at 18 to <24 weeks since dose among adults aged ≥65 years, albeit with confidence intervals overlapping that of Nunes et al. Link-Gelles reported VE against hospitalisation up to 43 weeks since dose for any XBB vaccine in the VISION Network in the United States [31]. Among immunocompetent adults aged ≥18 years, VE against COVID-19 hospitalisation during XBB and JN.1 predominant periods was 21% (95% CI: 10; 31) at 17 to 25.5 weeks since dose. At 26 to 43 weeks, XBB-adapted vaccines no longer provided significant protection against hospitalisation (VE: −8% [95% CI: −19; 3]) [31]. We found a higher VE of 59.2% (95% CI: 43.9; 70.3) at a comparable intermediate timepoint since dose at 18 to <24 weeks. However, minimal to no remaining protection was apparent in either study at the latest time points, >26 or >30 weeks since dose. Together, these findings suggest XBB vaccines may provide limited protection against COVID-19 hospitalisation in the latter 6 months of a once-yearly vaccination schedule.

As shown by Shrestha et al. (2024), Tartof et al. (2024), and Caffrey et al. (2024), the effectiveness of the BNT162b2 XBB vaccine was higher during XBB predominance compared to its effectiveness during JN.1 predominance [9–11]. This has also been shown in studies not limited to the BNT162b2 vaccine [35, 36]. VE tends to decrease when the circulating SARS-CoV-2 strains exhibit significant divergence from the subvariant targeted in the vaccine, supporting the need for periodic updates to variant-adapted COVID-19 vaccines. Nevertheless, we observed that BNT162b2 XBB vaccine was effective against JN.1-associated hospitalisation for up to six months.

Our main findings remained consistent across various sensitivity analyses, including modifying the number and location of knots for symptom onset date and age and stratifying by prior COVID-19 infection status. Similar to our first analysis [12], adjusting for influenza vaccination status in the last 12 months increased VE estimates. The inclusion of influenza vaccination status as a covariate in adjusted models can reduce potential residual bias related to the likelihood of enrolment as a test-negative control [37].

The inclusion of multiple sites across four European countries was a strength of this multi-centre study. This enabled broader surveillance to identify the JN.1 subvariant early, allowing follow-up for up to 46 weeks after XBB vaccination. We determined vaccination status using vaccine registries, vaccination cards, and health records, which likely reduced misclassification of exposure. Patients’ case status was verified using PCR which reduced the likelihood of misclassifying cases and controls. Furthermore, utilising the test-negative approach and requiring both cases and controls to be hospitalised with a SARI illness helped to account for unmeasured confounding factors linked to healthcare-seeking behaviours and exposure to respiratory pathogens.

Our study is subject to several limitations. Although results from sensitivity analyses were similar to the main results, our study nonetheless could be subject to residual confounding, particularly related to factors that are difficult to accurately measure, such as history of COVID-19 and time since the last infection. While we did collect information on prior infection status, this information was frequently missing, and prior infections are likely underestimated, given known discrepancies in the underreporting of SARS-CoV-2 infections [38]. Overall, 71% of the patients with SARI included in our analysis did not have a documented prior SARS-CoV-2 infection. Second, limited sample size for some stratified analyses prevented VE estimation or reduced the precision of VE estimated for certain in-hospital endpoints, such as the use of respiratory support. There was a high proportion (505/2025; 25%) of missing data for in-hospital respiratory support, which reduced the statistical power. Missingness was most prevalent in patients who did not receive the BNT162b2 XBB vaccine and was equally distributed among unvaccinated test-positive cases and test-negative controls (both 34%). Third, the generalisability of our findings beyond European populations primarily comprised of older adults is unknown.

We found that the effectiveness of the BNT162b2 XBB vaccine waned over time, with no evidence of remaining protection against hospitalisation by about 30 weeks since dose. These findings have important implications for the frequency of COVID-19 vaccination for high-risk patients, such as those with comorbid conditions and older adults. As evidenced by the July–August 2024 summer increase in COVID-19 hospitalisations in Europe, COVID-19 activity remains unpredictable, unlike the seasonal patterns displayed by influenza and some other respiratory viruses. Our findings support the need for additional vaccination in the spring season to maintain year-round protection against constantly changing circulating SARS-CoV-2 strains and unexpected increases in COVID-19 activity.

## Statements

### Ethical statement

Ethical approval for this study was obtained from Ethisch Comité UZA; Comitato Etico Regionale della Liguria (Genoa); Comité Ético de Investigación con Medicamentos del Departamento de Salud Arnau de Vilanova (Valencia); Comité Ético de Investigación con Medicamentos del Hospital Universitari Vall d’Hebron (Barcelona); Ethikkommission des Fachbereichs Medizin der Goethe-Universität (Frankfurt); Ethikkommission Ulm.

## Data Availability

De-identified data that underlie the results reported in this article (text, tables, figures, and appendices) may be obtained in accordance with id.DRIVE data access policy.

## Acknowledgements

The authors thank all current and former partners of COVIDRIVE and id.DRIVE (AstraZeneca, Bavarian Nordic, CureVac, FISABIO, GSK, Janssen, Novavax, Moderna, Pfizer, Sanofi, THL, Valneva, and P95) for their contribution to the Partnership. The authors thank Prof. Elizabeth Miller, a member of the id.DRIVE Independent Scientific Committee, for her valuable scientific advice to the id.DRIVE studies. The authors thank Pfizer, Novavax, and Valneva for their funding contribution to the data collection of the study reported in this publication.

The authors acknowledge the contribution of Sultan Abduljawad and Shanti Pather of BioNTech.

The authors thank Hospital Universitario Vall d’Hebrón, Hospital Universitari Germans Trias i Pujol, University Hospital Ulm, Centro Interuniversitario per la Ricerca sull’Influenza e le altre Infezioni Trasmissibili, Universitair Ziekenhuis Antwerpen, Le Centre Hospitalier Universitaire St Pierre, Valencia Hospital Network for the Study of Infectious Diseases (VAHNSI)-Fundación para el Fomento de la Investigación Sanitaria y Biomédica de la Comunitat Valenciana (Fisabio – Public Health), and University Hospital Frankfurt for collecting data of the study reported in this publication.

The authors wish to acknowledge the contribution of Griet Rebry (study operations), Juan Hernandez (statistician), Somsuvro Basu (medical writer) (all from P95 Clinical and Epidemiology Services), Brenda Marquina Sánchez (coordinator) of FISABIO, and all other members of the id.DRIVE Co-Coordination team.

## Authors’ contributions

This study was designed by members of the id.DRIVE consortium in collaboration with Pfizer. AA, IC, BG, GI, GLtK, C Martin, AM-I, AOS, SO-R, and GR are principal investigators at the study sites and were responsible for data collection and site coordination. Study site principal investigators gathered the data in collaboration with P95 and Pfizer. LD is involved in the coordination of the id.DRIVE study. All statistical analysis was performed by TMPT and EC-S. The data were interpreted by HRV, LdM, JLN, MM, C Marques, LC, SV, JY, LJ, JMM, and KB. LdM and LD accessed and verified the underlying data in the manuscript. HRV and LdM wrote the first draft of the manuscript. All authors reviewed, provided feedback on the manuscript drafts, and approved the manuscript for submission.

