## Supplemental Materials for "Durability of the BNT162b2 XBB.1.5-adapted vaccine against JN.1 hospitalisation in Europe, October 2023 to August 2024: A test-negative case-control study using the id.DRIVE platform"

#### Main supplementary tables and figures (arranged as they appear in the main manuscript)

**Supplementary Table S1. Start dates of the analysis period, per study site**

| Country | Study sites | Number of hospitals | Prospective data collection start date | XBB end | Transition period (XBB to JN.1) | JN.1 start |
|---|---|---|---|---|---|---|
| **Italy**[1] | Inter-university Research Centre on Influenza and Other Transmissible Infections (CIRI-IT) | 6 | October 2, 2023 | December 3, 2023 | December 4, 2023 to January 1, 2024 | January 2, 2024 |
| **Germany**[2] | University Hospital Frankfurt | 1 | October 2, 2023 | November 11, 2023 | November 12 to December 31 2023 | January 1, 2024 |
| | University Hospital Ulm | 1 | October 2, 2023 | | | |
| **Belgium**[3] | Universitair Ziekenhuis Antwerpen | 1 | December 1, 2023 | November 15, 2023 | November 16 to December 29 2023 | December 30, 2023 |
| | Le Centre Hospitalier Universitaire St Pierre, Brussels | 1 | December 1, 2023 | | | |
| **Spain**[4] | Valencia Hospital Network for the Study of Infectious Diseases (VAHNSI), Valencia | 5 | December 1, 2023 (retrospective data collection start date: October 2, 2023) | November 5, 2023 | November 6 to December 3 2023 | December 4, 2023 |
| | Ospital Universitari Germans Trias I Pujol , Badalona, Barcelona | 1 | December 1, 2023 | | | |
| | Hospital Universitari Vall d'Hebron, Barcelona | 1 | December 1, 2023 | | | |

Sources:
[1]https://www.epicentro.iss.it/coronavirus/bollettino/Bollettino-sorveglianza-integrata-COVID-19_24-gennaio-2024.pdf
[2]https://public.data.rki.de/t/public/views/IGS_Dashboard/DashboardSublineages?%3Aembed=y&%3AisGuestRedirectFromVizportal=y
[3] https://lookerstudio.google.com/embed/reporting/c14a5cfc-cab7-4812-848c-0369173148ab/page/urrUC
[4]https://www.isciii.es/QueHacemos/Servicios/VigilanciaSaludPublicaRENAVE/EnfermedadesTransmisibles/Paginas/Temporada_Gripe_23-24.aspx

**Supplementary Table S2. Chronic conditions and risk factors: diagnostic codes[1] and definitions**

| Covariate | Definition |
|---|---|
| **Chronic conditions** | |
| Asthma | • **Any of the following diagnostic codes (International Classification of Diseases, 10th revision [ICD-10]);** https://icd.who.int/browse10/2019/en**):** J45, J46<br>• **INCLUDING**: predominantly allergic asthma, nonallergic asthma, status asthmaticus, acute severe asthma<br>• **EXCLUDING:** chronic asthmatic (obstructive) bronchitis, chronic obstructive asthma, eosinophilic asthma, lung diseases due to external agents |
| Lung disease | • **Any of the following diagnostic codes (ICD-10):** A15-16, A19, A31.0, B33.4, E84.0, J40-44, J47, J60-70, J80-84, J85-86, J90-91, J92.0, J93-94, J95-99<br>• **INCLUDING:** tuberculosis (pulmonary, miliary, but not that of other systems), atypical mycobacteria, cystic fibrosis, chronic obstructive pulmonary disease (COPD), bronchiectasis and other chronic sequelae of infections, chronic lung diseases due to external agents, interstitial lung diseases, pleural diseases, respiratory failure<br>• **EXCLUDING:** acute respiratory infections, lung cancer, diseases of pulmonary circulation, pleural plaques without asbestos, previous uncomplicated pneumothorax |
| Cardiovascular disease | • **Any of the following diagnostic codes (ICD-10):** A52.0, B37.6, I01-02, I05-09, I11.0, I13.0, I13.2, I20-25, I26-28, I30-43, I44- 46, I48, I49.0, I49.5, I50-52, I70-71, Q20-Q28<br>• **INCLUDING:** all conditions of heart and large vessels that are chronic or likely to have chronic sequelae, cardiovascular syphilis, endo-, myo- and pericarditis, rheumatic fever, chronic rheumatic heart diseases, congenital malformations, hypertensive (renal) diseases with heart failure, ischaemic heart diseases, diseases of pulmonary circulation, atherosclerosis, cardiomyopathies, most conduction disorders, heart failure, aortic aneurysms & dissection, other heart diseases and their complications<br>• **EXCLUDING:** uncomplicated hypertension, previous uncomplicated pulmonary embolism (with no lasting cardiac insufficiency), paroxysmal tachycardias, most cases of premature depolarisation. |
| Hypertension | • **Any of the following diagnostic codes (ICD-10):** I10, I11.9, I12, I13.1, I13.9, I15<br>• **INCLUDING**: essential (primary) hypertension, secondary hypertension<br>• **EXCLUDING:** hypertensive heart/renal disease with (congestive) heart failure |
| Chronic liver disease | • **Any of the following diagnostic codes (ICD-10):** B18, B19, K70, K71.1, K71.3, K71.4-9, K72.1, K72.9, K73, K74, K75.2-4, K75.8, K75.9, K76.0-2, K76.5, K76.6, K76.9<br>• **INCLUDING:** all conditions of the liver that are chronic or likely to have chronic sequelae<br>• **EXCLUDING:** acute liver disease, liver cancer/metastasis, liver transplant |
| Renal disease | • **Any of the following diagnostic codes (ICD-10):** N18-19<br>• **INCLUDING:** decreased kidney function shown by glomerular filtration rate (GFR) of less than 60 mL/min per 1·73 $m^2$, or markers of kidney damage*, or both, of at least 3 months duration, regardless of the underlying cause.<br>***Markers of kidney damage include albuminuria (30 mg/24 hours; Albumin: Creatinine ratio > 3 mg/mmol), urine sediment abnormalities, electrolyte, and other abnormalities due to tubular disorders, abnormalities detected by histology, structural abnormalities detected by imaging, and a history of kidney transplantation (Based on KDIGO and NICE guidelines).* |

| Covariate | Definition |
|---|---|
| | • **EXCLUDING:** acute kidney failure, clinically nonsignificant kidney cysts |
| Diabetes type II | • **Any of the following diagnostic codes (ICD-10):** E11<br>• **INCLUDING:** Diabetes mellitus type II (adult-onset, maturity-onset, obese and non-obese, non-insulin-dependent diabetes of the young)<br>• **EXCLUDING:** malnutrition-related diabetes mellitus, diabetes mellitus in pregnancy or puerperium, glycosuria, impaired glucose tolerance, postsurgical hypoinsulinaemia |
| Cancer | • **Any of the following diagnostic codes (ICD-10):** C00-97, D37-48, Z85, Z92.3, Z92.6<br>• **INCLUDING:** all malignant neoplasms (both solid and haematologic) with potential to metastasise, either in treatment, active follow-up, or <5 years post curative treatment<br>• **EXCLUDING:** benign & in situ neoplasms. Basal cell carcinomas. Any cancer previously treated with curative intent & in complete remission for ≥5 years. |
| Immunodeficiency (or organ transplant) | • **Any of the following diagnostic codes (ICD-10):** B20-B24, D80–84, D89, Z94<br>• **INCLUDING:**<br>**Solid organ transplant**<br>**Haematopoietic stem cell transplantation**<br>**Primary immunodeficiency**<br>**Advanced or untreated HIV infection**: people with HIV and CD4 cell counts less than 200/mm3, history of an AIDS-defining illness without immune reconstitution, or clinical manifestations of symptomatic HIV)<br>**Iatrogenic immunodeficiencies**: systemic treatment of more than two weeks duration, in the three months preceding symptom onset, with any of the following: corticosteroid (≥20 mg prednisolone daily or equivalent when administered for 2 or more weeks), ciclosporin, tacrolimus, mycophenolate, methotrexate, azathioprine, tumour necrosis factor alpha (TNF-α) blockers and other biological or cytostatic drugs with immunosuppressive effect. The treatment causing immunodeficiency has to be ongoing or expected to still cause immunodeficiency (treatment-dependent durations).<br>The option 'Iatrogenic immunodeficiency' is to be selected when the treatment responsible for the immunodeficiency is prescribed for a condition already identified in this questionnaire (e.g., haematopoietic stem cell transplantation or cancer)<br>• **EXCLUDING:** disorders of the immune system which do not lead to immunosuppression (e.g. some autoimmune conditions), adequately treated HIV infection. |

[1] *The list of codes has been developed based on the following sources:*

a) https://www.cdc.gov/coronavirus/2019-ncov/hcp/clinical-care/underlyingconditions.html
b) https://www.ecdc.europa.eu/en/publications-data/core-protocol-ecdc-studies-covid-19-vaccine-effectiveness-against-0
c) https://www.ecdc.europa.eu/en/seasonal-influenza/prevention-and-control/vaccines/risk-groups
d) https://www.cdc.gov/rsv/high-risk/older-adults.html
e) https://archive.cdc.gov/#/details?url=https://www.cdc.gov/nchs/nhis/tobacco/tobacco_glossary.htm

**Supplementary Figure S1. Flow diagram of the study population**

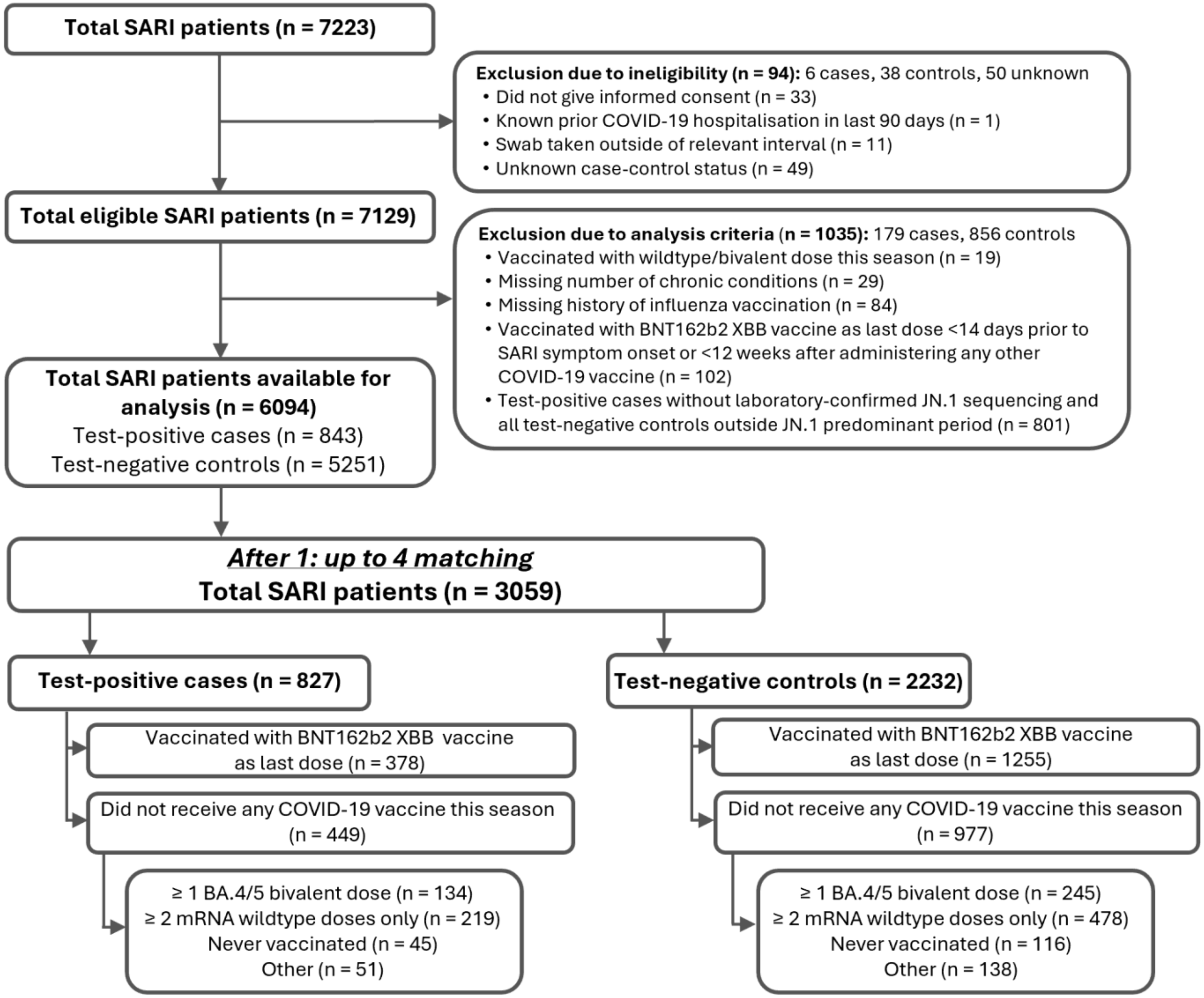

*Abbreviations: n, number; SARI, severe acute respiratory infections.*

**Supplementary Table S3. Characteristics of study participants according to receipt of BNT162b2 XBB vaccine and SARS-CoV-2 case classification during JN.1 predominant period, with prior vaccination categories as reference groups.**

| | Total, N (col %) | Received BNT162b2 XBB vaccine in 2023–2024 season, (N, col %) | | Received ≥ 1 BA.4/5 bivalent dose, (N, col %) | |
|---|---|---|---|---|---|
| | | Test-positive cases | Test-negative controls | Test-positive cases | Test-negative controls |
| **Total** | 1879 | 377 | 1145 | 133 | 224 |
| **Sex** | | | | | |
| Female | 869 (46.2) | 165 (43.8) | 543 (47.4) | 64 (48.1) | 97 (43.3) |
| Male | 1010 (53.8) | 212 (56.2) | 602 (52.6) | 69 (51.9) | 127 (56.7) |
| **Age, years** | | | | | |
| Median (IQR) | 80.0 (72.0, 87.0) | 81.0 (73.0, 88.0) | 80.0 (73.0, 88.0) | 76.0 (66.0, 85.0) | 77.0 (68.0, 85.0) |
| 18-49 | 38 (2.0) | 3 (0.8) | 17 (1.5) | 10 (7.5) | 8 (3.6) |
| 50-64 | 193 (10.3) | 32 (8.5) | 104 (9.1) | 21 (15.8) | 36 (16.1) |
| ≥65 | 1648 (87.7) | 342 (90.7) | 1024 (89.4) | 102 (76.7) | 180 (80.4) |
| **Body mass index (BMI)[1],** kg/m$^2$ | | | | | |
| Median (IQR) | 26.0 (23.1, 29.7) | 25.8 (22.8, 29.1) | 26.1 (23.1, 30.0) | 25.8 (23.5, 28.9) | 26.1 (23.1, 30.4) |
| Underweight (BMI <18.5) | 52 (3.5) | 15 (4.9) | 22 (2.4) | 4 (4.1) | 11 (7.3) |
| Normal weight (BMI ≥18.5 to <25.0) | 551 (37.3) | 114 (37.4) | 353 (38.2) | 36 (36.7) | 48 (32.0) |
| Overweight (BMI ≥25.0 to <30.0) | 490 (33.2) | 108 (35.4) | 304 (32.9) | 36 (36.7) | 42 (28.0) |
| Obese (BMI ≥30.0)[2] | 385 (26.0) | 68 (22.3) | 246 (26.6) | 22 (22.4) | 49 (32.7) |
| Missing | 401 | 72 | 220 | 35 | 74 |
| **Number of chronic conditions** | | | | | |
| 0-1[3] | 452 (24.1) | 102 (27.1) | 260 (22.7) | 34 (25.6) | 56 (25.0) |
| 2 | 539 (28.7) | 95 (25.2) | 350 (30.6) | 34 (25.6) | 60 (26.8) |
| ≥3 | 888 (47.3) | 180 (47.7) | 535 (46.7) | 65 (48.9) | 108 (48.2) |
| **Type of chronic condition** | | | | | |

| | | | | | |
|---|---|---|---|---|---|
| Asthma | 157 (8.4) | 26 (6.9) | 99 (8.6) | 14 (10.5) | 18 (8.0) |
| Lung disease | 738 (39.3) | 124 (32.9) | 464 (40.5) | 48 (36.1) | 102 (45.5) |
| Cardiovascular disease | 891 (47.4) | 170 (45.1) | 558 (48.7) | 54 (40.6) | 109 (48.7) |
| Hypertension | 1321 (70.3) | 258 (68.4) | 828 (72.3) | 87 (65.4) | 148 (66.1) |
| Renal disease | 117 (6.2) | 25 (6.6) | 59 (5.2) | 14 (10.5) | 19 (8.5) |
| Liver disease | 351 (18.7) | 89 (23.6) | 194 (16.9) | 28 (21.1) | 40 (17.9) |
| Type 2 diabetes | 654 (34.8) | 137 (36.3) | 407 (35.5) | 46 (34.6) | 64 (28.6) |
| Immunodeficiency or cancer[4] | 402 (21.4) | 93 (24.7) | 207 (18.1) | 39 (29.3) | 63 (28.1) |
| **Reported prior SARS-CoV-2 infection[1]** | | | | | |
| Yes | 580 (35.5) | 123 (36.1) | 358 (35.4) | 37 (34.3) | 62 (35.8) |
| No | 1052 (64.5) | 218 (63.9) | 652 (64.6) | 71 (65.7) | 111 (64.2) |
| Missing | 247 | 36 | 135 | 25 | 51 |
| **Influenza vaccination within 12 months prior to hospitalisation** | | | | | |
| Yes | 1566 (83.3) | 357 (94.7) | 1088 (95.0) | 50 (37.6) | 71 (31.7) |
| No | 313 (16.7) | 20 (5.3) | 57 (5.0) | 83 (62.4) | 153 (68.3) |
| **Symptom onset, month** | | | | | |
| Dec 2023 | 459 (24.4) | 59 (15.6) | 289 (25.2) | 40 (30.1) | 71 (31.7) |
| Jan 2024 | 426 (22.7) | 59 (15.6) | 291 (25.4) | 31 (23.3) | 45 (20.1) |
| Feb 2024 | 105 (5.6) | 14 (3.7) | 73 (6.4) | 5 (3.8) | 13 (5.8) |
| Mar 2024 | 60 (3.2) | 8 (2.1) | 46 (4.0) | 4 (3.0) | 2 (0.9) |
| Apr 2024 | 55 (2.9) | 10 (2.7) | 38 (3.3) | 1 (0.8) | 6 (2.7) |
| May 2024 | 131 (7.0) | 21 (5.6) | 102 (8.9) | 4 (3.0) | 4 (1.8) |
| Jun 2024 | 254 (13.5) | 71 (18.8) | 132 (11.5) | 17 (12.8) | 34 (15.2) |
| Jul 2024 | 253 (13.5) | 92 (24.4) | 108 (9.4) | 24 (18.0) | 29 (12.9) |
| Aug 2024 | 136 (7.2) | 43 (11.4) | 66 (5.8) | 7 (5.3) | 20 (8.9) |
| **Country of hospitalisation** | | | | | |
| Belgium | 5 (0.3) | 1 (0.3) | 4 (0.3) | 0 (0.0) | 0 (0.0) |
| Germany | 0 (0.0) | 0 (0.0) | 0 (0.0) | 0 (0.0) | 0 (0.0) |
| Italy | 74 (3.9) | 14 (3.7) | 34 (3.0) | 7 (5.3) | 19 (8.5) |
| Spain | 1800 (95.8) | 362 (96.0) | 1107 (96.7) | 126 (94.7) | 205 (91.5) |

| | | | | | |
|---|---|---|---|---|---|
| **Length of hospital stay, days** | | | | | |
| Median (IQR) | 6.0 (4.0, 9.0) | 5.0 (4.0, 8.0) | 6.0 (4.0, 9.0) | 6.0 (4.0, 9.0) | 6.0 (4.0, 10.0) |
| 1–3 | 379 (20.2) | 83 (22.0) | 226 (19.7) | 27 (20.3) | 43 (19.2) |
| 4–6 | 690 (36.7) | 158 (41.9) | 397 (34.7) | 49 (36.8) | 86 (38.4) |
| 7–13 | 581 (30.9) | 99 (26.3) | 389 (34.0) | 35 (26.3) | 58 (25.9) |
| 14–27 | 184 (9.8) | 29 (7.7) | 112 (9.8) | 17 (12.8) | 26 (11.6) |
| 28–41 | 32 (1.7) | 7 (1.9) | 13 (1.1) | 4 (3.0) | 8 (3.6) |
| ≥42 | 13 (0.7) | 1 (0.3) | 8 (0.7) | 1 (0.8) | 3 (1.3) |
| **Level of respiratory support[1,5]** | | | | | |
| None | 326 (21.6) | 69 (23.9) | 199 (20.5) | 28 (30.4) | 30 (19.1) |
| Oxygen therapy | 991 (65.6) | 186 (64.4) | 643 (66.1) | 58 (63.0) | 104 (66.2) |
| Non-invasive ventilation | 185 (12.2) | 34 (11.8) | 124 (12.7) | 5 (5.4) | 22 (14.0) |
| Invasive mechanical ventilation | 8 (0.5) | 0 (0.0) | 7 (0.7) | 0 (0.0) | 1 (0.6) |
| ECMO | 1 (0.1) | 0 (0.0) | 0 (0.0) | 1 (1.1) | 0 (0.0) |
| Missing | 368 | 88 | 172 | 41 | 67 |
| **Level of SARI severity[1]** | | | | | |
| Hospital admission without intensive care unit (ICU) admission and without in-hospital death | 1708 (91.0) | 346 (92.0) | 1039 (90.7) | 125 (94.0) | 198 (88.8) |
| ICU admission without in-hospital death | 35 (1.9) | 4 (1.1) | 24 (2.1) | 1 (0.8) | 6 (2.7) |
| In-hospital death | 134 (7.1) | 26 (6.9) | 82 (7.2) | 7 (5.3) | 19 (8.5) |
| Missing | 2 | 1 | 0 | 0 | 1 |

| | | **Received BNT162b2 XBB vaccine in 2023–2024 season, (N, col %)** | | **Received ≥ 2 wild type doses only, (N, col %)** | |
|---|---|---|---|---|---|
| | **Total, N (col %)** | **Test-positive cases** | **Test-negative controls** | **Test-positive cases** | **Test-negative controls** |
| **Total** | 2146 | 378 | 1104 | 218 | 446 |
| **Sex** | | | | | |
| Female | 1008 (47.0) | 165 (43.7) | 534 (48.4) | 115 (52.8) | 194 (43.5) |

| | | | | | |
|---|---|---|---|---|---|
| Male | 1138 (53.0) | 213 (56.3) | 570 (51.6) | 103 (47.2) | 252 (56.5) |
| **Age, years** | | | | | |
| Median (IQR) | 79.0 (69.0, 86.0) | 81.0 (73.0, 88.0) | 80.5 (73.0, 88.0) | 77.0 (58.0, 85.0) | 69.0 (52.0, 80.0) |
| 18-49 | 153 (7.1) | 3 (0.8) | 17 (1.5) | 31 (14.2) | 102 (22.9) |
| 50-64 | 270 (12.6) | 32 (8.5) | 100 (9.1) | 47 (21.6) | 91 (20.4) |
| ≥65 | 1723 (80.3) | 343 (90.7) | 987 (89.4) | 140 (64.2) | 253 (56.7) |
| **Body mass index (BMI)[1],** kg/m$^2$ | | | | | |
| Median (IQR) | 25.8 (22.8, 29.4) | 25.8 (22.8, 29.1) | 26.0 (23.1, 30.0) | 24.9 (22.6, 29.4) | 25.1 (22.1, 28.5) |
| Underweight (BMI <18.5) | 64 (3.8) | 15 (4.9) | 28 (3.1) | 10 (6.3) | 11 (3.4) |
| Normal weight (BMI ≥18.5 to <25.0) | 658 (39.0) | 115 (37.6) | 340 (37.9) | 68 (43.0) | 135 (41.4) |
| Overweight (BMI ≥25.0 to <30.0) | 548 (32.5) | 108 (35.3) | 290 (32.3) | 38 (24.1) | 112 (34.4) |
| Obese (BMI ≥30.0)[2] | 417 (24.7) | 68 (22.2) | 239 (26.6) | 42 (26.6) | 68 (20.9) |
| Missing | 459 | 72 | 207 | 60 | 120 |
| **Number of chronic conditions** | | | | | |
| 0-1[3] | 645 (30.1) | 103 (27.2) | 248 (22.5) | 81 (37.2) | 213 (47.8) |
| 2 | 615 (28.7) | 95 (25.1) | 358 (32.4) | 58 (26.6) | 104 (23.3) |
| ≥3 | 886 (41.3) | 180 (47.6) | 498 (45.1) | 79 (36.2) | 129 (28.9) |
| **Type of chronic condition** | | | | | |
| Asthma | 172 (8.0) | 26 (6.9) | 94 (8.5) | 13 (6.0) | 39 (8.7) |
| Lung disease | 754 (35.1) | 125 (33.1) | 452 (40.9) | 47 (21.6) | 130 (29.1) |
| Cardiovascular disease | 927 (43.2) | 170 (45.0) | 527 (47.7) | 93 (42.7) | 137 (30.7) |
| Hypertension | 1384 (64.5) | 258 (68.3) | 799 (72.4) | 126 (57.8) | 201 (45.1) |
| Renal disease | 123 (5.7) | 25 (6.6) | 56 (5.1) | 19 (8.7) | 23 (5.2) |
| Liver disease | 366 (17.1) | 89 (23.5) | 178 (16.1) | 45 (20.6) | 54 (12.1) |
| Type 2 diabetes | 675 (31.5) | 137 (36.2) | 388 (35.1) | 60 (27.5) | 90 (20.2) |
| Immunodeficiency or cancer[4] | 441 (20.5) | 93 (24.6) | 200 (18.1) | 59 (27.1) | 89 (20.0) |
| **Reported prior SARS-CoV-2 infection[1]** | | | | | |
| Yes | 668 (35.6) | 124 (36.3) | 344 (35.2) | 64 (35.0) | 136 (36.5) |
| No | 1206 (64.4) | 218 (63.7) | 632 (64.8) | 119 (65.0) | 237 (63.5) |

| | | | | | |
|---|---|---|---|---|---|
| Missing | 272 | 36 | 128 | 35 | 73 |
| **Influenza vaccination within 12 months prior to hospitalisation** | | | | | |
| Yes | 1548 (72.1) | 358 (94.7) | 1048 (94.9) | 55 (25.2) | 87 (19.5) |
| No | 598 (27.9) | 20 (5.3) | 56 (5.1) | 163 (74.8) | 359 (80.5) |
| **Symptom onset, month** | | | | | |
| Nov 2023 | 3 (0.1) | 0 (0.0) | 0 (0.0) | 3 (1.4) | 0 (0.0) |
| Dec 2023 | 465 (21.7) | 59 (15.6) | 261 (23.6) | 35 (16.1) | 110 (24.7) |
| Jan 2024 | 476 (22.2) | 59 (15.6) | 273 (24.7) | 50 (22.9) | 94 (21.1) |
| Feb 2024 | 136 (6.3) | 14 (3.7) | 66 (6.0) | 16 (7.3) | 40 (9.0) |
| Mar 2024 | 80 (3.7) | 8 (2.1) | 46 (4.2) | 6 (2.8) | 20 (4.5) |
| Apr 2024 | 65 (3.0) | 10 (2.6) | 36 (3.3) | 3 (1.4) | 16 (3.6) |
| May 2024 | 166 (7.7) | 21 (5.6) | 123 (11.1) | 10 (4.6) | 12 (2.7) |
| Jun 2024 | 288 (13.4) | 71 (18.8) | 125 (11.3) | 39 (17.9) | 53 (11.9) |
| Jul 2024 | 304 (14.2) | 92 (24.3) | 108 (9.8) | 40 (18.3) | 64 (14.3) |
| Aug 2024 | 163 (7.6) | 44 (11.6) | 66 (6.0) | 16 (7.3) | 37 (8.3) |
| **Country of hospitalisation** | | | | | |
| Belgium | 5 (0.2) | 1 (0.3) | 4 (0.4) | 0 (0.0) | 0 (0.0) |
| Germany | 14 (0.7) | 0 (0.0) | 0 (0.0) | 5 (2.3) | 9 (2.0) |
| Italy | 192 (8.9) | 15 (4.0) | 29 (2.6) | 37 (17.0) | 111 (24.9) |
| Spain | 1935 (90.2) | 362 (95.8) | 1071 (97.0) | 176 (80.7) | 326 (73.1) |
| **Length of hospital stay, days** | | | | | |
| Median (IQR) | 6.0 (4.0, 9.0) | 5.0 (4.0, 8.0) | 6.0 (4.0, 9.0) | 6.0 (4.0, 10.0) | 6.0 (4.0, 10.0) |
| 1–3 | 448 (20.9) | 83 (22.0) | 221 (20.0) | 45 (20.6) | 99 (22.2) |
| 4–6 | 773 (36.0) | 158 (41.8) | 395 (35.8) | 78 (35.8) | 142 (31.8) |
| 7–13 | 663 (30.9) | 99 (26.2) | 369 (33.4) | 55 (25.2) | 140 (31.4) |
| 14–27 | 209 (9.7) | 30 (7.9) | 100 (9.1) | 34 (15.6) | 45 (10.1) |
| 28–41 | 33 (1.5) | 7 (1.9) | 13 (1.2) | 3 (1.4) | 10 (2.2) |
| ≥42 | 20 (0.9) | 1 (0.3) | 6 (0.5) | 3 (1.4) | 10 (2.2) |
| **Level of respiratory support[1,5]** | | | | | |
| None | 415 (24.2) | 69 (23.8) | 178 (18.8) | 44 (29.3) | 124 (37.6) |

| | | | | | |
|---|---|---|---|---|---|
| Oxygen therapy | 1083 (63.1) | 187 (64.5) | 633 (67.0) | 94 (62.7) | 169 (51.2) |
| Non-invasive ventilation | 204 (11.9) | 34 (11.7) | 126 (13.3) | 10 (6.7) | 34 (10.3) |
| Invasive mechanical ventilation | 13 (0.8) | 0 (0.0) | 8 (0.8) | 2 (1.3) | 3 (0.9) |
| ECMO | 0 (0.0) | 0 (0.0) | 0 (0.0) | 0 (0.0) | 0 (0.0) |
| Missing | 431 | 88 | 159 | 68 | 116 |
| **Level of SARI severity**[1] | | | | | |
| Hospital admission without intensive care unit (ICU) admission and without in-hospital death | 1975 (92.2) | 347 (92.0) | 1007 (91.3) | 201 (93.1) | 420 (94.2) |
| ICU admission without in-hospital death | 47 (2.2) | 4 (1.1) | 23 (2.1) | 7 (3.2) | 13 (2.9) |
| In-hospital death | 120 (5.6) | 26 (6.9) | 73 (6.6) | 8 (3.7) | 13 (2.9) |
| Missing | 4 | 1 | 1 | 2 | 0 |

| | | **Received BNT162b2 XBB vaccine in 2023–2024 season, (N, col %)** | | **Never vaccinated, (N, col %)** | |
|---|---|---|---|---|---|
| | **Total, N (col %)** | **Test-positive cases** | **Test-negative controls** | **Test-positive cases** | **Test-negative controls** |
| **Total** | 1521 | 377 | 1001 | 44 | 99 |
| **Sex** | | | | | |
| Female | 710 (46.7) | 165 (43.8) | 474 (47.4) | 19 (43.2) | 52 (52.5) |
| Male | 811 (53.3) | 212 (56.2) | 527 (52.6) | 25 (56.8) | 47 (47.5) |
| **Age, years** | | | | | |
| Median (IQR) | 80.0 (71.0, 88.0) | 81.0 (73.0, 88.0) | 80.0 (73.0, 88.0) | 69.0 (55.5, 82.2) | 66.0 (46.0, 80.0) |
| 18-49 | 56 (3.7) | 3 (0.8) | 17 (1.7) | 6 (13.6) | 30 (30.3) |
| 50-64 | 151 (9.9) | 32 (8.5) | 91 (9.1) | 11 (25.0) | 17 (17.2) |
| ≥65 | 1314 (86.4) | 342 (90.7) | 893 (89.2) | 27 (61.4) | 52 (52.5) |
| **Body mass index (BMI)[1],** kg/m$^2$ | | | | | |
| Median (IQR) | 25.8 (22.9, 29.4) | 25.8 (22.8, 29.1) | 25.9 (23.0, 30.0) | 23.9 (20.8, 26.5) | 25.0 (22.8, 28.8) |
| Underweight (BMI <18.5) | 44 (3.6) | 15 (4.9) | 21 (2.6) | 4 (13.3) | 4 (5.6) |

| | | | | | |
|---|---|---|---|---|---|
| Normal weight (BMI ≥18.5 to <25.0) | 478 (39.2) | 114 (37.4) | 318 (39.2) | 14 (46.7) | 32 (44.4) |
| Overweight (BMI ≥25.0 to <30.0) | 391 (32.1) | 108 (35.4) | 256 (31.5) | 6 (20.0) | 21 (29.2) |
| Obese (BMI ≥30.0)[2] | 306 (25.1) | 68 (22.3) | 217 (26.7) | 6 (20.0) | 15 (20.8) |
| Missing | 302 | 72 | 189 | 14 | 27 |
| **Number of chronic conditions** | | | | | |
| 0-1[3] | 412 (27.1) | 102 (27.1) | 235 (23.5) | 21 (47.7) | 54 (54.5) |
| 2 | 422 (27.7) | 95 (25.2) | 307 (30.7) | 5 (11.4) | 15 (15.2) |
| ≥3 | 687 (45.2) | 180 (47.7) | 459 (45.9) | 18 (40.9) | 30 (30.3) |
| **Type of chronic condition** | | | | | |
| Asthma | 133 (8.7) | 26 (6.9) | 87 (8.7) | 4 (9.1) | 16 (16.2) |
| Lung disease | 582 (38.3) | 124 (32.9) | 410 (41.0) | 17 (38.6) | 31 (31.3) |
| Cardiovascular disease | 697 (45.8) | 170 (45.1) | 486 (48.6) | 18 (40.9) | 23 (23.2) |
| Hypertension | 1045 (68.7) | 258 (68.4) | 718 (71.7) | 22 (50.0) | 47 (47.5) |
| Renal disease | 88 (5.8) | 25 (6.6) | 51 (5.1) | 5 (11.4) | 7 (7.1) |
| Liver disease | 256 (16.8) | 89 (23.6) | 158 (15.8) | 3 (6.8) | 6 (6.1) |
| Type 2 diabetes | 516 (33.9) | 137 (36.3) | 346 (34.6) | 9 (20.5) | 24 (24.2) |
| Immunodeficiency or cancer[4] | 282 (18.5) | 93 (24.7) | 169 (16.9) | 9 (20.5) | 11 (11.1) |
| **Reported prior SARS-CoV-2 infection[1]** | | | | | |
| Yes | 464 (34.2) | 123 (36.1) | 310 (34.8) | 4 (10.0) | 27 (31.8) |
| No | 893 (65.8) | 218 (63.9) | 581 (65.2) | 36 (90.0) | 58 (68.2) |
| Missing | 164 | 36 | 110 | 4 | 14 |
| **Influenza vaccination within 12 months prior to hospitalisation** | | | | | |
| Yes | 1315 (86.5) | 357 (94.7) | 950 (94.9) | 3 (6.8) | 5 (5.1) |
| No | 206 (13.5) | 20 (5.3) | 51 (5.1) | 41 (93.2) | 94 (94.9) |
| **Symptom onset, month** | | | | | |
| Nov 2023 | 0 (0.0) | 0 (0.0) | 0 (0.0) | 0 (0.0) | 0 (0.0) |
| Dec 2023 | 332 (21.8) | 59 (15.6) | 233 (23.3) | 11 (25.0) | 29 (29.3) |
| Jan 2024 | 326 (21.4) | 59 (15.6) | 242 (24.2) | 8 (18.2) | 17 (17.2) |
| Feb 2024 | 80 (5.3) | 14 (3.7) | 60 (6.0) | 2 (4.5) | 4 (4.0) |

| | | | | | |
|---|---|---|---|---|---|
| Mar 2024 | 50 (3.3) | 8 (2.1) | 35 (3.5) | 2 (4.5) | 5 (5.1) |
| Apr 2024 | 53 (3.5) | 10 (2.7) | 40 (4.0) | 1 (2.3) | 2 (2.0) |
| May 2024 | 118 (7.8) | 21 (5.6) | 88 (8.8) | 1 (2.3) | 8 (8.1) |
| Jun 2024 | 215 (14.1) | 71 (18.8) | 129 (12.9) | 5 (11.4) | 10 (10.1) |
| Jul 2024 | 224 (14.7) | 92 (24.4) | 108 (10.8) | 12 (27.3) | 12 (12.1) |
| Aug 2024 | 123 (8.1) | 43 (11.4) | 66 (6.6) | 2 (4.5) | 12 (12.1) |
| **Country of hospitalisation** | | | | | |
| Belgium | 5 (0.3) | 1 (0.3) | 4 (0.4) | 0 (0.0) | 0 (0.0) |
| Germany | 0 (0.0) | 0 (0.0) | 0 (0.0) | 0 (0.0) | 0 (0.0) |
| Italy | 55 (3.6) | 14 (3.7) | 34 (3.4) | 2 (4.5) | 5 (5.1) |
| Spain | 1461 (96.1) | 362 (96.0) | 963 (96.2) | 42 (95.5) | 94 (94.9) |
| **Length of hospital stay, days** | | | | | |
| Median (IQR) | 6.0 (4.0, 9.0) | 5.0 (4.0, 8.0) | 6.0 (4.0, 9.0) | 5.0 (4.0, 7.2) | 5.0 (3.0, 8.0) |
| 1–3 | 321 (21.1) | 83 (22.0) | 199 (19.9) | 10 (22.7) | 29 (29.3) |
| 4–6 | 568 (37.3) | 158 (41.9) | 358 (35.8) | 18 (40.9) | 34 (34.3) |
| 7–13 | 474 (31.2) | 99 (26.3) | 339 (33.9) | 10 (22.7) | 26 (26.3) |
| 14–27 | 129 (8.5) | 29 (7.7) | 88 (8.8) | 5 (11.4) | 7 (7.1) |
| 28–41 | 22 (1.4) | 7 (1.9) | 13 (1.3) | 1 (2.3) | 1 (1.0) |
| ≥42 | 7 (0.5) | 1 (0.3) | 4 (0.4) | 0 (0.0) | 2 (2.0) |
| **Level of respiratory support[1,5]** | | | | | |
| None | 251 (20.2) | 69 (23.9) | 164 (19.3) | 6 (20.0) | 12 (16.0) |
| Oxygen therapy | 820 (65.9) | 186 (64.4) | 563 (66.2) | 20 (66.7) | 51 (68.0) |
| Non-invasive ventilation | 164 (13.2) | 34 (11.8) | 117 (13.8) | 3 (10.0) | 10 (13.3) |
| Invasive mechanical ventilation | 9 (0.7) | 0 (0.0) | 6 (0.7) | 1 (3.3) | 2 (2.7) |
| ECMO | 0 (0.0) | 0 (0.0) | 0 (0.0) | 0 (0.0) | 0 (0.0) |
| Missing | 277 | 88 | 151 | 14 | 24 |
| **Level of SARI severity[1]** | | | | | |
| Hospital admission without intensive care unit (ICU) admission and without in-hospital death | 1377 (90.6) | 346 (92.0) | 913 (91.2) | 33 (75.0) | 85 (85.9) |

| | | | | | |
|---|---|---|---|---|---|
| ICU admission without in-hospital death | 40 (2.6) | 4 (1.1) | 26 (2.6) | 4 (9.1) | 6 (6.1) |
| In-hospital death | 103 (6.8) | 26 (6.9) | 62 (6.2) | 7 (15.9) | 8 (8.1) |
| Missing | 1 | 1 | 0 | 0 | 0 |

*Abbreviations: ECMO, extracorporeal membrane oxygenation; ICU, intensive care unit; IQR, interquartile range; N, number.*

*[1] Percentages exclude subjects with missing values.*

*[2] Obese classification was based on both BMI ≥30.0 kg/m$^2$ and obesity as a categorical variable (yes/no).*

*[3] The categories "0" and "1" among the number of chronic conditions are counted as separate categories of number of chronic conditions, based on the list of chronic conditions provided in Supplementary Table S2. Here, the cell counts among participants having "0–1" chronic conditions are provided as a composite.*

*[4] The categories "Cancer" and "Immunodeficiency" among chronic conditions are identified as separate types of chronic conditions. Here, the cell counts among participants having "Immunodeficiency or cancer" are provided as a composite.*

*[5] Respiratory support level, presented in ascending order of severity with the highest level used during hospital stay determine the category, are mutually exclusive: 1) none; 2) oxygen therapy (e.g., nasal cannula or mask); 3) non-invasive ventilation (support without endotracheal intubation such as high flow nasal oxygen, continuous or bi-level positive airway pressure); 4) invasive mechanical ventilation (support with endotracheal intubation); and 5) extracorporeal membrane oxygenation (ECMO).*

**Supplementary Figure S2. Vaccine effectiveness against JN.1-related hospitalisation in SARI patients who received at least one dose of BNT162b2 XBB vaccine using various prior vaccination categories as comparison groups.**

| Exposure | Total, N | Test-positive cases: N (%) | Test-negative controls: N(%) | Days since last vaccine dose: Median (Q1; Q3) | VE (95% CI)[1] |
|---|---|---|---|---|---|
| **Overall** | | | | | |
| BNT162b2 XBB vaccine | 1633 | 378 (23%) | 1255 (77%) | 107 (65; 223) | 47.9% (37.0; 56.9) |
| Did not receive any COVID-19 vaccine this season | 1426 | 449 (31%) | 977 (69%) | 744 (566; 898)[2] | |
| **Stratification of comparison groups[3]** | | | | | |
| BNT162b2 XBB vaccine | 1522 | 377 (25%) | 1145 (75%) | 108 (64; 227) | 48.6% (38.9; 56.7) |
| ≥1 BA.4/5 bivalent dose | 357 | 133 (37%) | 224 (63%) | 449 (405; 577)[2] | |
| BNT162b2 XBB vaccine | 1482 | 378 (26%) | 1104 (74%) | 120 (67; 228) | 46.4% (37.1; 54.4) |
| ≥2 mRNA wild type doses only | 664 | 218 (33%) | 446 (67%) | 826 (738; 943)[2] | |
| BNT162b2 XBB vaccine | 1378 | 377 (27%) | 1001 (73%) | 132 (68; 233) | 26.7% (-38.8; 61.3) |
| Never vaccinated | 143 | 44 (31%) | 99 (69%) | | |

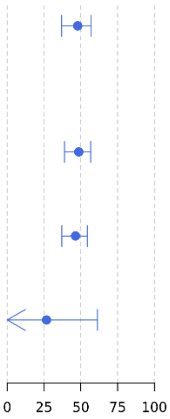

*Abbreviations: N, number; SARI, severe acute respiratory infections.*

[1] *Vaccine effectiveness (VE) estimates are adjusted for date of symptom onset, age, sex, number of chronic conditions and receipt of influenza vaccine 12 months prior to current hospital admission.*
[2] *'Never vaccinated' subjects were excluded when calculating the median (interquartile range) of time since last vaccine dose among patients who did not receive any dose of a COVID-19 vaccine in the 2023–2024 autumn/winter season.*
[3] *Stratifications of comparison groups were assessed on matched data per exposure of interest and numbers will therefore not add up to the numbers provided in Table 1.*

**Supplementary Figure S3. Vaccine effectiveness against JN.1-related hospitalisation in SARI patients who received at least one dose of BNT162b2 XBB vaccine compared to patients who did not receive any dose of a COVID-19 vaccine in the 2023–2024 autumn/winter with durability stratified by hospital severity level.**

| Strata | Exposure | Total, N | Test-positive cases: N (%) | Test-negative controls: N (%) | Days since last vaccine dose: Median (Q1; Q3) | VE (95% CI)[1] |
|---|---|---|---|---|---|---|
| **Hospital admission only** | | | | | | |
| Overall | Yes | 1489 | 347 (23%) | 1142 (77%) | 110 (66; 223) | 46.5% (31.9; 58.0) |
| | No | 1304 | 408 (31%) | 896 (69%) | 747 (566; 898)[2] | |
| **Weeks since BNT162b2 XBB vaccine dose** | | | | | | |
| 2 to <30 wks | Yes | 1043 | 176 (17%) | 867 (83%) | 78 (56; 124) | 64.1% (55.6; 70.9) |
| | No | 1304 | 408 (31%) | 896 (69%) | 747 (566; 898)[2] | |
| 2 to <12 wks | Yes | 590 | 94 (16%) | 496 (84%) | 59 (45; 71) | 68.4% (54.9; 77.9) |
| | No | 1304 | 408 (31%) | 896 (69%) | 747 (566; 898)[2] | |
| 12 to <18 wks | Yes | 196 | 25 (13%) | 171 (87%) | 96 (89; 107) | 76.1% (69.3; 81.3) |
| | No | 1304 | 408 (31%) | 896 (69%) | 747 (566; 898)[2] | |
| 18 to <24 wks | Yes | 92 | 20 (22%) | 72 (78%) | 144 (137; 154) | 53.3% (36.0; 66.0) |
| | No | 1304 | 408 (31%) | 896 (69%) | 747 (566; 898)[2] | |
| 24 to <30 wks | Yes | 165 | 37 (22%) | 128 (78%) | 195 (182; 203) | 51.8% (40.9; 60.6) |
| | No | 1304 | 408 (31%) | 896 (69%) | 747 (566; 898)[2] | |
| 30 to <46 wks | Yes | 446 | 171 (38%) | 275 (62%) | 248 (230; 272) | 2.1% (-43.0; 32.9) |
| | No | 1304 | 408 (31%) | 896 (69%) | 747 (566; 898)[2] | |
| **ICU admission/in-hospital death** | | | | | | |
| Overall | Yes | 142 | 30 (21%) | 112 (79%) | 86 (62; 216) | 55.4% (21.3; 74.8) |
| | No | 118 | 39 (33%) | 79 (67%) | 716 (555; 900)[2] | |
| **Weeks since BNT162b2 XBB vaccine dose** | | | | | | |
| 2 to <30 wks | Yes | 102 | 16 (16%) | 86 (84%) | 71 (55; 98) | 70.7% (51.8; 82.2) |
| | No | 118 | 39 (33%) | 79 (67%) | 716 (555; 900)[2] | |
| 2 to <12 wks | Yes | 69 | 10 (14%) | 59 (86%) | 62 (42; 71) | 70.3% (37.2; 86.0) |
| | No | 118 | 39 (33%) | 79 (67%) | 716 (555; 900)[2] | |
| 12 to <18 wks | Yes | 16 | 3 (19%) | 13 (81%) | 98 (88; 104) | 59.3% (5.6; 82.5) |
| | No | 118 | 39 (33%) | 79 (67%) | 716 (555; 900)[2] | |
| 18 to <24 wks | Yes | 9 | 0 (0%) | 9 (100%) | 151 (140; 166) | |
| | No | 118 | 39 (33%) | 79 (67%) | 716 (555; 900)[2] | |
| 24 to <30 wks[3] | Yes | 8 | 3 (38%) | 5 (62%) | 192 (190; 201) | |
| | No | 118 | 39 (33%) | 79 (67%) | 716 (555; 900)[2] | |
| 30 to <46 wks[3] | Yes | 40 | 14 (35%) | 26 (65%) | 254 (238; 282) | |
| | No | 118 | 39 (33%) | 79 (67%) | 716 (555; 900)[2] | |

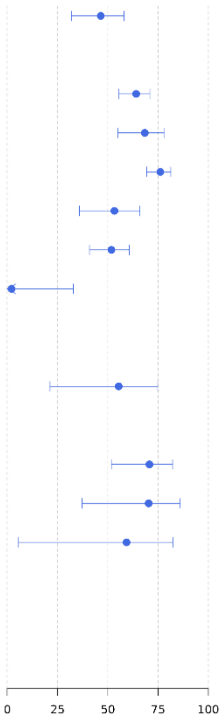

*Abbreviations: ICU, intensive care unit; N, number; SARI, severe acute respiratory infections.*

*[1] Vaccine effectiveness (VE) estimates are adjusted for date of symptom onset, age, sex, number of chronic conditions and receipt of influenza vaccine 12 months prior to current hospital admission.*
*[2] 'Never vaccinated' subjects were excluded when calculating the median (interquartile range) of time since last vaccine dose in the unexposed group (patients who did not receive any dose of a COVID-19 vaccine in the 2023–2024 autumn/winter season).*
*[3] Results not shown due to unreliable interpretation of the results because of limited number of subjects in the strata and wide 95% CIs.*

### Appendix

#### A.1. General framework for the COVID-19 vaccine effectiveness analysis

To address potential intra-cluster correlation among subjects from the same Study Contributor, Generalized Estimating Equations (GEE) was employed for analysing subject-level data. In this context, a Study Contributor was considered as a distinct cluster. Heterogeneity between Study Contributors may exist due to the differences in recruitment, local differences in the intensity of the epidemic and healthcare practices.

##### Analysis overview

The COVID-19 vaccine effectiveness (VE)) against hospitalisation due to laboratory-confirmed COVID-19 disease was estimated as:

$$VE = (1 - OR) \text{ x } 100\%$$

where OR denotes the odds ratio, which is calculated as the ratio of the odds of vaccination among SARS-CoV-2 test-positive cases to the odds of vaccination among SARS-CoV-2 test-negative controls.

Let $K$ denote the number of clusters (Study Contributors), $n_k$ represent the number of included patients from Study Contributor $k\,(k = 1, \ldots, K)$ (from now will be referred to as cluster). We denote $Y_{ik}$ the variable representing the infection status of subject $i$ from cluster $k$, $Y_{ik} = 1$ [if positive-case] and $Y_{ik} = 0$ [if negative-control]. $X_1$ is the variable representing the vaccination status of subject $i$ from cluster $k$, $X_{1ik} = 1$ [if vaccinated] and $X_{1ik} = 0$ [if unvaccinated or other exposure definition depending on the analyses].

Assuming that $Y_{ik}$ follow a binomial distribution with $\pi_{ik} = P(Y_{ik} = 1 | X_{1ik})$, the probability of being a COVID-19 positive case by laboratory-confirmed RT-PCR test or other equivalent tests (from now on, we will refer to it as a COVID-19 positive case). Conditionally on the vaccination status and using a logit-function, we can relate:

$$\text{logit}\,(\pi_{ik}) = \log\left(\frac{\pi_{ik}}{1 - \pi_{ik}}\right) = \beta_0 + \beta_1 X_{1ik} \quad [1]$$

Hence, the odds of having COVID-19 in the vaccinated group (i.e. $X_1 = 1$) is $\pi_{vaccinated}/(1 - \pi_{vaccinated})$ = exp($\beta_0 + \beta_1$) while the odds of having COVID-19 in the reference group (for example, unvaccinated this season, i.e., $X_1 = 0$) is $\pi_{reference}/(1 - \pi_{reference})$ = exp($\beta_0$). Consequently, the odds ratio (OR) of having COVID-19 in the vaccinated group compared to the reference group is estimated as:

$$OR = \frac{\pi_{vaccinated}/(1-\pi_{vaccinated})}{\pi_{reference}/(1-\pi_{reference})} = \frac{\exp(\beta_0 + \beta_1)}{\exp(\beta_0)} = \exp(\beta_1).$$

Finally, due to the symmetrical characteristic of the OR, the VE is calculated as follows:

$$VE = \left(1 - \exp(\beta_1)\right) \times 100\%$$

The GEE approach was used to obtain the population-averaged estimate (over Study Contributors) of parameter of interest (in this case, $\beta_1$), adjusting for other covariates (potential confounders or effect modifiers), depending on the analyses. The VE estimates with their 95% CIs were reported. We computed 95% Wald CI, assuming normal sampling distributions of the estimates. To accomplish the interpretation of the outputs, we report the median and interquartile range (IQR) of time since last dose (measured in days) for the exposed subjects contributing to the estimate.

#### Generalized Estimating Equations (GEE) approach

*Overview of the approach*

Generalized Estimating Equations (GEE), initially proposed by Liang *et al.* and Zeger *et al.* [1, 2], is a widely used approach for analysing clustered data. GEE employs a marginal logistic regression model to address potential intra-cluster correlation, particularly in cases where outcomes of subjects from a cluster (Study Contributor in our study) may exhibit greater similarity compared to subjects from other Study Contributors. Treating a Study Contributor as a cluster, with the cluster size equal to the number of subjects from the corresponding Study Contributor included in the analysis, GEE models provide population-averaged estimates. Notably, GEE avoids the need to specify the full joint likelihood function while performing model fitting, a challenge in cases of discrete response variables. Hence, this approach requires only the specification of the first two marginal moments – the mean vector and the variance-covariance matrix.

In the GEE approach, the robust sandwich estimator, which relies on a working correlation matrix, yields a consistent and asymptotically normal estimator [2]. A working correlation matrix is a matrix presenting an assumed correlation structure between observations from the same cluster. The clusters are assumed to be independent of each other. If the assumed working correlation structure is close to the truth, some efficiency might be gained. However, the estimates from GEE are consistent even if the working correlation structure is mis-specified. Exchangeable, independent, and first-order autoregressive structures are examples of commonly used working correlation structures [1].

*Model fitting procedure*

For all our analyses, we assumed an independent working correlation among outcomes of subjects within the same cluster. Seaman *et al.* and Kahan *et al*. [3, 4] have shown that using exchangeable working correlation structure might lead to biased estimates in case there are cluster-based confounding (CBC) and/or informative cluster sizes (ICS) present and hence, an independent working correlation structure is recommended. GEE models were fitted using the PGEE package in R [5]. The PGEE package implements a penalised GEE procedure, as proposed by Wang *et al*. [6], designed for the analysis of correlated data with high-dimensional covariates. This method accommodates situations with the diverging number of parameters to be estimated. In our analyses, however, we slightly modified the written package to allow for a "ridge" penalty term. The inclusion of the *ridge* penalty term is useful to handle model fitting difficulty when there is multicollinearity present [7]. Multicollinearity might present when there is a strong correlation among predictor variables, which might lead to unstable and biased standard errors [8]. Moreover, this approach also provides robust estimates with regard to the choice of the number and location of knots (for symptom onset date [and age]).

GEE routines typically obtain initial coefficient estimates from corresponding generalised linear models (GLM) and update these parameters iteratively until convergence. However, in the presence of separation, utilising a regular (i.e., unpenalised) GLM routine can result in infinite estimates of the odds ratio (on the log scale). To circumvent these issues, the initial coefficients are obtained from ridge regression (i.e., a GLM with a ridge penalty) using the *glmnet* package [9], and these coefficients are subsequently utilised in the PGEE call. The use of *glmnet* also facilitates obtaining an approximate estimate of the penalty parameter used in the penalised GEE model. This penalty parameter is selected by minimising out-of-sample binomial deviance through *five-fold cross-validation*, employing the *one-standard error* rule (using the *glmnet* package [10]). The initial coefficient associated with the penalty parameter determined by the one-standard error rule is chosen as the coefficient for the PGEE fitting procedure.

To ensure reproducibility in the cross-validation procedure, the seed number 23 is employed by calling set.seed (23) before the *glmnet* function is called. The variables passed to the *glmnet* call are *not* necessary to be standardised (i.e., not rescaled to have 0 mean and unit variance). This is because the function itself already implements data processing procedures to ensure that the included covariates (either categorical variables or B-spline bases of the continuous variables) are on the same scale. Finally, to guarantee reliable VE estimates, the GEE analysis was only conducted if there are a minimum of 2 subjects in each cell of the 2x2 table (exposure x outcome) without considering stratification for other covariates and a minimum total sample size of 50 subjects.

*Confounder adjusted VE estimate*

The **confounder-adjusted VE estimate** was obtained from a logistic regression model that includes the following covariates: *vaccination status*, *symptom onset date*, *age*, *sex*, the *number of chronic conditions*, and the receipt of influenza vaccination 12 months prior to symptom onset. The reason that the symptom onset was included in the model was to adjust for changes in the SARS-CoV-2 incidence as well as time-varying vaccination coverage which made it a potential confounder. As the study duration increases, the complexity of the effect for onset date will need to increase. The effects of symptom onset date and subjects‘ ages were modelled using (penalised) cubic regression splines. The number of chronic conditions was treated as a categorical variable, taking values 0 (no chronic condition present), 1 (only one chronic condition present), 2 (two chronic conditions present), or ≥3 (at least three chronic conditions present). Finally, the receipt of influenza vaccination was treated as a binary variable, taking value 1 if ever vaccinated with influenza vaccine(s) 12 months prior to symptom onset and 0, otherwise.

The effects of symptom onset date ($X_2$), subject's age in years ($X_3$), subject's gender assigned at birth ($X_4$), the number of chronic conditions ($X_5$) and the receipt of influenza vaccination were included into the GEE models. Formula [1] is extended to:

$$\text{logit}\,(\pi_{ik}) = \beta_0 + \beta_1 X_{1ik} + s_2(X_{2ik}) + s_3(X_{3ik}) + \beta_4 X_{4ik} + \beta_5 X_{5ik} + \beta_6 X_{6ik} \quad [1a]$$

Here, $X_2$ and $X_3$ are treated as continuous variables. $s_2$ and $s_3$ are (cubic spline) smooth functions of the variable presenting symptom onset date ($X_2$) and subjects' age ($X_3$), respectively, with subscripts $i, k$ defined above. The use of a smooth function allows capturing a flexible relationship between the odds of having COVID-19 and symptom onset date as well as subject's age. For symptom onset date, we assigned a sufficiently large number to the number of degrees of freedom of the spline term. Since most of the subjects enrolled in the study between the end of November 2023 till August 2024, it is reasonable to fit the model with 9 equally-sized knots (with the distance between knots to be approximately 1 month) for the cubic spline capturing symptom onset date effect. This assumption led to a cubic spline term with 12 degrees of freedom. For the effect of age, we performed the analysis using 2 knots (at age 50 and 65) for its spline term, leading to the number of degrees of freedom equal to 5. If there was less than 10% of subjects who were younger than 50 years old, we performed the analysis using only 1 knot (at age 65) instead of the spline term of age, leading to a total number of degrees of freedom equal to 4.

Several sensitivity analyses with respect to the number and location of knots for both symptom onset date and age variables were conducted. The results showed that the proposed method is robust against the different choices of number and location of knots.

#### Missing data

When a subject lacks data on outcome, exposure, or specific covariates (dependent on the model), that specific subject was excluded from the analysis. This method, referred to as complete case analysis (CCA), uses only subjects with complete information for all variables included in the analysis. When data is missing completely at random (MCAR), CCA yields unbiased estimates. Under the assumption that missingness happens completely at random (Missing Completely At Random – MCAR mechanism), the set of complete observations is considered a random sample from the population. Thus, the CCA will not produce biased estimates [11].

Given that the COVID-19 status is part of the primary data collection, it was anticipated that case status should be available for nearly all subjects. However, data on exposure status and, in particular, potential confounders might be missing for a portion of subjects. These details were typically obtained from pre-existing medical records, vaccine registries, etc., which existed before the SARI episode. Consequently, it was reasonable to assume that the MCAR mechanism holds. However, the CCA approach, which utilises a subset of the whole sample when missingness occurs, leads to a loss of precision, especially when the number of subjects with missing data is relatively high. Consequently, if the CCA makes use of a dataset with more than 10% loss in sample size in comparison with the full dataset, we would implement multiple imputation techniques. The subsequent analysis would then be carried out on the imputed datasets. However, in our analysis, there was limited number of subjects excluded due to missingness. As a consequence, no multiple imputation was performed.

#### A.2. Sample size calculations, technical specifications

In general, the goal of the sample size calculations is to determine the minimum sample size necessary to ensure desirable properties of the VE estimates. Simulation-based methods to determine sample sizes are developed to closely mimic the actual study design and proposed analytical approaches at the cost of requiring additional parameter assumptions and computational burden.

#### Simulation-based approach

##### <u>Data generation workflow</u>

###### Notation

Before describing the data generation workflow, the following parameters which act as input for the model have to be defined:

- $VE_{x,overall}$: the overall VE of exposure x, the corresponding odds ratio is $OR_{x,overall} = 1 - \frac{VE_{x,overall}}{100}$.
- $c = P(unexposed|control)$: proportion of unexposed subjects among the controls
- $P_x = P(exposure\ x|exposed, control)$: brand share of exposure x among the exposed
- $r$: ratio of cases to control (that is, number of cases per one control)

###### General set-up

In each simulation run, a dataset was constructed by combining data generated for a number of individual sites. We denote the total number of study sites as $k$ and the total sample size as $N$. Additionally, it was assumed that each site contributed the same number of subjects ($\frac{N}{k}$). In order to allow for variability in the underlying vaccine effects across study sites, the VE can be different from site to site. In the next section, we described how data for one site was generated given the study site-specific VEs for all exposures. The subsequent section describes how the VE were varied across the study sites to introduce between-site variability.

###### Simulating data at the site level

For each site, $\frac{N}{k} \times \frac{r}{1+r}$ cases and $\frac{N}{k} \times \frac{1}{1+r}$ controls were simulated.

Vaccine exposure status for the controls was generated from a multinomial distribution with the probability of being unexposed equal to $c$ and the probability of being exposed to brand $x$ equal to $(1 - c)P_x$ (with $\sum_x P_x = 1$).

For each $\frac{N}{k} \times \frac{r}{1+r}$ of the cases, the vaccine exposure status was then generated from a multinomial distribution with the probability of being unexposed $P(unexposed|case) = \frac{1}{1+\sum_x \mathrm{OR}_x * \frac{(1-c)P_x}{c}}$ and the probability of being exposed to brand $x$ equal to $P(exposure\ x|case) = \mathrm{OR}_x * \frac{(1-c)P_x}{c} * P(unexposed|case)$, where

$$\frac{\frac{P(exposure\ x|case)}{P(unexposed|case)}}{\frac{P(exposure\ x|control)}{P(unexposed|control)}} = OR_{x,site}$$

To incorporate the expected between-site heterogeneity, for each study site, a site-specific odds ratio ($OR_{x,site}$) was generated from a log-normal distribution with a median of $1 - \frac{VE_{x,overall}}{100}$ and variance on the log scale of 0.05. The value of the variance parameter on the log scale was selected to be 0.05 as it introduced an amount of between-site heterogeneity and was in line with the heterogeneity seen in a previous database study [1]. The expected value of the VE over the sites was then equal to $100 \times \left(1 - \exp\left(\log\left(1 - \frac{VE_{x,overall}}{100}\right) + \frac{0.05}{2}\right)\right)$.

#### Estimates

For each simulated dataset, an estimate of the VE and the corresponding 95% CI is obtained using the generalized estimating equations (GEE) method:

- The expected OR on the log-scale of the treatment effect was estimated using a logistic regression model with the disease status as the outcome and the exposure as a covariate. The estimates were obtained using the GEE method in which the sites were considered clusters and the variances were calculated using a robust sandwich estimator.
- The estimated log OR and the corresponding CI were then back-transformed to obtain an estimate of the overall VE and its 95% CI.
- The overall VE estimate and the length of the CI were stored for each simulation.

A total of 100 to 200 simulations were performed, which led to stable Monte Carlo CIs while limiting the computational burden.

The expected range of the 95% CI is defined as the mean range of the CI obtained from all simulations. For each measure, 95% Monte Carlo CIs were constructed based on the respective Monte Carlo standard errors observed in the simulations.

#### Sample size calculation for the study reported in the manuscript

The sample size of the number of SARI patients in the autumn/winter 2023-2024 season required to obtain a VE estimate of the primary objective with an expected width of the 95% CI fulfilling a prespecified expectation relies on the following assumptions: An overall vaccination coverage of 40%, a proportion among vaccinated subjects vaccinated with at least one dose of BNT162b2 XBB.1.5-adapted mRNA COVID-19 vaccine (Pfizer/BioNTech 2023–2024 formulation, hereafter referred to as the BNT162b2 XBB vaccine) in the autumn/winter 2023-2024 season of 90%, a VE of 70% for all other vaccine brands and an anticipated VE of 70% for BNT162b2 XBB vaccine, a control-case ratio of 4:1, assuming 10 Study Contributors and a GEE analysis. 10 Study Contributors were the number of Study Contributors anticipated to take part in the study at the beginning.

We calculated a required sample size of at least 410 SARI patients. Since this calculation was based on crude VE estimates (without covariates adjustment), we conservatively inflated the sample size by a factor of 1.2 to account for adjusted estimates. Therefore, the final targeted sample size is 410 x 1.2 = 492 SARI patients (including (410-82) x 1.2 = 394 controls and 82 x 1.2 = 98 COVID-19 cases). This sample size is expected to yield an adjusted VE estimate with a 95% CI width of ≤50%.

#### Sample size taking into account matching cases and controls

The matching process is anticipated to result in a reduction of the sample size for analysis by approximately 40% based on observations from the data in the autumn/winter 2022-2023 season (hospital admission between 02 October 2022 – 31 May 2023), given a matching ratio of 1 (positive) case to a maximum of 4 (negative) controls. Consequently, to achieve a sample size of 492 SARI patients for the analysis of the primary VE, 492 x 5/3 = 820 SARI patients would be required.

#### A.3. Matching cases and controls

Up to four test-negative controls were matched to each case based on the following variables:

- Study Contributor (and thus, Country)
- Temporal proximity of symptom onset (2 weeks)

The matching process was conducted in R using an exact matching methodology. The number of controls matched to each case were examined. Matching was done separately for each vaccine effectiveness analysis. For example, in the analysis of the primary objective, only cases and controls that fit the definitions of "*vaccinated with at least one dose of* BNT162b2 XBB vaccine *in the 2023-2024 autumn/winter seasons"* or "*not vaccinated this season*" were matched.

If more than 4 controls meet the matching criteria, controls would be selected based on the closeness of symptom onset dates to the case, prioritising shorter time gaps. Ultimately, for the primary objective analysis, 16 test-positive cases were excluded due to a lack of matched test-negative controls, and 3019 negative controls were removed because the case-to-control ratio was 1:6.

#### A.4. Sensitivity analysis results

In this section, we are providing results from various sensitivity analyses for all VE reported in Figure 1 in the main manuscript.

##### Sensitivity analysis with regard to location and number of knots of symptom onset dates

**Supplementary Figure S4 [Sensitivity analysis 1a]** Vaccine effectiveness against JN.1-related hospitalisation in SARI patients who received at least one dose of BNT162b2 XBB vaccine compared to patients who did not receive any dose of a COVID-19 vaccine in the 2023–2024 autumn/winter season. The location of knots for symptom onset dates were chosen at 15 December 2023, 15 January 2024, 15 February 2024, 15 March 2024, 15 April 2024, 15 May 2024, 15 June 2024, 15 July 2024, and 15 August 2024.

| Strata | Exposure | Total, N | Test-positive cases: N (%) | Test-negative controls: N (%) | Days since last vaccine dose: Median (Q1; Q3) | VE (95% CI)[1] |
|---|---|---|---|---|---|---|
| **Overall** | | | | | | |
| Overall | Yes | 1633 | 378 (23%) | 1255 (77%) | 107 (65; 223) | 48.4% (42.8; 53.4) |
| | No | 1426 | 449 (31%) | 977 (69%) | 744 (566; 898)[2] | |
| **Age group (years)** | | | | | | |
| 18-64y | Yes | 166 | 35 (21%) | 131 (79%) | 106 (62; 226) | 66.4% (55.3; 74.8) |
| | No | 525 | 153 (29%) | 372 (71%) | 791 (701; 916)[2] | |
| ≥65y | Yes | 1467 | 343 (23%) | 1124 (77%) | 107 (65; 223) | 44.9% (38.7; 50.5) |
| | No | 901 | 296 (33%) | 605 (67%) | 717 (510; 846)[2] | |
| 65-79y | Yes | 589 | 129 (22%) | 460 (78%) | 115 (60; 220) | 33.4% (-10.3; 59.7) |
| | No | 457 | 132 (29%) | 325 (71%) | 724 (533; 873)[2] | |
| ≥80y | Yes | 878 | 214 (24%) | 664 (76%) | 104 (68; 224) | 53.2% (44.1; 60.8) |
| | No | 444 | 164 (37%) | 280 (63%) | 692 (476; 836)[2] | |
| **Type of chronic condition** | | | | | | |
| Asthma | Yes | 139 | 26 (19%) | 113 (81%) | 86 (60; 204) | 48.6% (-41.4; 81.3) |
| | No | 130 | 37 (28%) | 93 (72%) | 737 (556; 898)[2] | |
| Lung disease | Yes | 649 | 125 (19%) | 524 (81%) | 99 (63; 216) | 44.6% (28.2; 57.2) |
| | No | 440 | 128 (29%) | 312 (71%) | 723 (496; 846)[2] | |
| Cardiovascular disease | Yes | 780 | 170 (22%) | 610 (78%) | 95 (63; 220) | 58.0% (43.3; 69.0) |
| | No | 503 | 179 (36%) | 324 (64%) | 723 (507; 845)[2] | |
| Hypertension | Yes | 1161 | 258 (22%) | 903 (78%) | 102 (64; 220) | 50.2% (37.2; 60.5) |
| | No | 747 | 258 (35%) | 489 (65%) | 724 (519; 848)[2] | |
| Chronic liver disease | Yes | 89 | 25 (28%) | 64 (72%) | 131 (66; 224) | 58.3% (37.5; 72.1) |
| | No | 107 | 46 (43%) | 61 (57%) | 719 (467; 878)[2] | |
| Renal disease | Yes | 297 | 89 (30%) | 208 (70%) | 99 (66; 226) | 45.2% (21.6; 61.7) |
| | No | 203 | 82 (40%) | 121 (60%) | 732 (554; 885)[2] | |
| Type 2 diabetes | Yes | 576 | 137 (24%) | 439 (76%) | 108 (67; 227) | 41.7% (29.5; 51.9) |
| | No | 355 | 123 (35%) | 232 (65%) | 718 (541; 882)[2] | |
| Immunodeficiency or cancer [3] | Yes | 316 | 93 (29%) | 223 (71%) | 144 (60; 233) | 53.2% (4.0; 77.2) |
| | No | 332 | 125 (38%) | 207 (62%) | 728 (534; 872)[2] | |
| **Number of chronic conditions** | | | | | | |
| 0-1[4] | Yes | 383 | 103 (27%) | 280 (73%) | 134 (68; 228) | 31.9% (18.5; 43.2) |
| | No | 583 | 160 (27%) | 423 (73%) | 775 (648; 926)[2] | |
| 2 | Yes | 488 | 95 (19%) | 393 (81%) | 107 (67; 221) | 69.8% (64.9; 74.1) |
| | No | 339 | 112 (33%) | 227 (67%) | 737 (543; 893)[2] | |
| ≥3 | Yes | 762 | 180 (24%) | 582 (76%) | 100 (63; 220) | 42.0% (32.9; 49.9) |
| | No | 504 | 177 (35%) | 327 (65%) | 710 (489; 834)[2] | |
| **Weeks since BNT162b2 XBB vaccine dose** | | | | | | |
| 2 to <30 wks | Yes | 1146 | 193 (17%) | 953 (83%) | 77 (55; 120) | 64.3% (56.5; 70.7) |
| | No | 1426 | 449 (31%) | 977 (69%) | 744 (566; 898)[2] | |
| 2 to <12 wks | Yes | 660 | 105 (16%) | 555 (84%) | 59 (45; 71) | 68.2% (55.8; 77.2) |
| | No | 1426 | 449 (31%) | 977 (69%) | 744 (566; 898)[2] | |
| 12 to <18 wks | Yes | 212 | 28 (13%) | 184 (87%) | 96 (89; 107) | 74.9% (68.0; 80.3) |
| | No | 1426 | 449 (31%) | 977 (69%) | 744 (566; 898)[2] | |
| 18 to <24 wks | Yes | 101 | 20 (20%) | 81 (80%) | 145 (137; 155) | 59.3% (43.6; 70.6) |
| | No | 1426 | 449 (31%) | 977 (69%) | 744 (566; 898)[2] | |
| 24 to <30 wks | Yes | 173 | 40 (23%) | 133 (77%) | 194 (182; 203) | 51.7% (38.2; 62.3) |
| | No | 1426 | 449 (31%) | 977 (69%) | 744 (566; 898)[2] | |
| 30 to <46 wks | Yes | 487 | 185 (38%) | 302 (62%) | 250 (230; 272) | 7.7% (-24.3; 31.4) |
| | No | 1426 | 449 (31%) | 977 (69%) | 744 (566; 898)[2] | |
| **Severity level[5]** | | | | | | |
| Hospital admission, no ICU, no IH death | Yes | 1489 | 347 (23%) | 1142 (77%) | 110 (66; 223) | 47.3% (39.8; 53.9) |
| | No | 1304 | 408 (31%) | 896 (69%) | 747 (566; 898)[2] | |
| ICU admission, no IH death | Yes | 29 | 4 (14%) | 25 (86%) | 77 (62; 215) | 63.6% (23.8; 82.7)* |
| | No | 50 | 15 (30%) | 35 (70%) | 778 (695; 882)[2] | |
| In-hospital death | Yes | 113 | 26 (23%) | 87 (77%) | 88 (62; 224) | 57.8% (-15.3; 84.5) |
| | No | 68 | 24 (35%) | 44 (65%) | 645 (525; 904)[2] | |
| ICU admission or IH death | Yes | 142 | 30 (21%) | 112 (79%) | 86 (62; 216) | 55.4% (21.3; 74.7) |
| | No | 118 | 39 (33%) | 79 (67%) | 716 (555; 900)[2] | |
| **Respiratory support level[6]** | | | | | | |
| None | Yes | 284 | 69 (24%) | 215 (76%) | 114 (62; 226) | 61.8% (45.6; 73.1) |
| | No | 308 | 98 (32%) | 210 (68%) | 778 (623; 918)[2] | |
| Oxygen therapy | Yes | 901 | 187 (21%) | 714 (79%) | 121 (68; 224) | 50.5% (31.5; 64.2) |
| | No | 596 | 187 (31%) | 409 (69%) | 737 (568; 899)[2] | |
| Non-invasive ventilation | Yes | 174 | 34 (20%) | 140 (80%) | 97 (61; 216) | -38.3% (-88.3; -1.5) |
| | No | 96 | 19 (20%) | 77 (80%) | 724 (479; 901)[2] | |
| Invasive mechanical ventilation | Yes | 9 | 0 (0%) | 9 (100%) | 69 (61; 97) | |
| | No | 10 | 3 (30%) | 7 (70%) | 722 (708; 786)[2] | |
| ECMO | Yes | 0 | 0 (0%) | 0 (0%) | | |
| | No | 1 | 1 (100%) | 0 (0%) | 519 (519; 519)[2] | |
| **Length of stay** | | | | | | |
| LOS ≥ 2 days | Yes | 1603 | 368 (23%) | 1235 (77%) | 107 (65; 223) | 49.6% (44.8; 53.9) |
| | No | 1379 | 436 (32%) | 943 (68%) | 744 (566; 899)[2] | |
| LOS ≥ 4 days | Yes | 1296 | 295 (23%) | 1001 (77%) | 102 (64; 222) | 50.5% (43.6; 56.6) |
| | No | 1113 | 359 (32%) | 754 (68%) | 744 (566; 898)[2] | |
| LOS ≥ 7 days | Yes | 692 | 137 (20%) | 555 (80%) | 88 (61; 210) | 61.5% (52.0; 69.2) |
| | No | 635 | 197 (31%) | 438 (69%) | 735 (563; 875)[2] | |

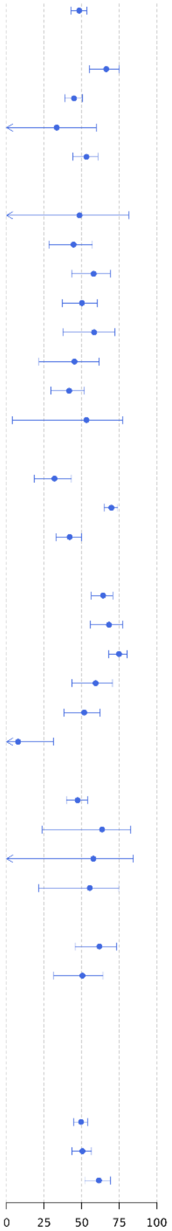

*Abbreviations: ECMO, extracorporeal membrane oxygenation; ICU, intensive care unit; IH, in-hospital; N, number; SARI, severe acute respiratory infections.*

[1] *Vaccine effectiveness (VE) estimates are adjusted for date of symptom onset, age, sex, number of chronic conditions and receipt of influenza vaccine in the 12 months prior to current hospital admission.*

[2] *'Never vaccinated' subjects were excluded when calculating the median (interquartile range) of time since last vaccine dose in the unexposed group (patients who did not receive any dose of a COVID-19 vaccine in the 2023–2024 autumn/winter season).*

[3] *The categories "Cancer" and "Immunodeficiency" among chronic conditions are counted as separate types of chronic conditions in all adjusted VE estimates. Here, the VE estimate among participants having "Immunodeficiency or cancer" is provided as a composite.*

[4] *The categories "0" and "1" among the number of chronic conditions are counted as separate categories of number of chronic conditions in all adjusted VE estimates. Here, the VE estimate among participants having "0–1"chronic conditions is provided as a composite.*

[5] *The categories "Hospital admission, no ICU, no IH death", "ICU admission, no IH death", and "In-hospital death" are mutually exclusive. An additional category is added to combine the latter two categories: "ICU admission or IH death".*

[6] *Respiratory support level, presented in ascending order of severity with the highest level used during hospital stay determine the category, are mutually exclusive: 1) none;, 2) oxygen therapy (e.g., nasal cannula or mask); 3) non-invasive ventilation (support without endotracheal intubation such as high flow nasal oxygen, continuous or bi-level positive airway pressure); 4) invasive mechanical ventilation (support with endotracheal intubation); and 5) extracorporeal membrane oxygenation.*

**Supplementary Figure S5 [Sensitivity analysis 1b]** Vaccine effectiveness against JN.1-related hospitalisation in SARI patients who received at least one dose of BNT162b2 XBB vaccine compared to patients who did not receive any dose of a COVID-19 vaccine in the 2023–2024 autumn/winter season. The location of knots for symptom onset dates were chosen at 1 January 2024, 15 February 2024, 01 April 2024, 15 May 2024, 01 July 2024, and 15 August 2024.

| Strata | Exposure | Total, N | Test-positive cases: N (%) | Test-negative controls: N (%) | Days since last vaccine dose: Median (Q1; Q3) | VE (95% CI)[1] |
|---|---|---|---|---|---|---|
| **Overall** | | | | | | |
| Overall | Yes | 1633 | 378 (23%) | 1255 (77%) | 107 (65; 223) | 47.9% (36.9; 57.0) |
| | No | 1426 | 449 (31%) | 977 (69%) | 744 (566; 898)[2] | |
| **Age group (years)** | | | | | | |
| 18-64y | Yes | 166 | 35 (21%) | 131 (79%) | 106 (62; 226) | 64.0% (54.5; 71.5) |
| | No | 525 | 153 (29%) | 372 (71%) | 791 (701; 916)[2] | |
| ≥65y | Yes | 1467 | 343 (23%) | 1124 (77%) | 107 (65; 223) | 45.1% (30.8; 56.5) |
| | No | 901 | 296 (33%) | 605 (67%) | 717 (510; 846)[2] | |
| 65-79y | Yes | 589 | 129 (22%) | 460 (78%) | 115 (60; 220) | 33.1% (-11.5; 59.9) |
| | No | 457 | 132 (29%) | 325 (71%) | 724 (533; 873)[2] | |
| ≥80y | Yes | 878 | 214 (24%) | 664 (76%) | 104 (68; 224) | 53.2% (43.9; 61.0) |
| | No | 444 | 164 (37%) | 280 (63%) | 692 (476; 836)[2] | |
| **Type of chronic condition** | | | | | | |
| Asthma | Yes | 139 | 26 (19%) | 113 (81%) | 86 (60; 204) | 47.2% (-44.7; 80.7) |
| | No | 130 | 37 (28%) | 93 (72%) | 737 (556; 898)[2] | |
| Lung disease | Yes | 649 | 125 (19%) | 524 (81%) | 99 (63; 216) | 44.5% (28.1; 57.2) |
| | No | 440 | 128 (29%) | 312 (71%) | 723 (496; 846)[2] | |
| Cardiovascular disease | Yes | 780 | 170 (22%) | 610 (78%) | 95 (63; 220) | 58.0% (42.9; 69.2) |
| | No | 503 | 179 (36%) | 324 (64%) | 723 (507; 845)[2] | |
| Hypertension | Yes | 1161 | 258 (22%) | 903 (78%) | 102 (64; 220) | 50.1% (28.8; 65.1) |
| | No | 747 | 258 (35%) | 489 (65%) | 724 (519; 848)[2] | |
| Chronic liver disease | Yes | 89 | 25 (28%) | 64 (72%) | 131 (66; 224) | 58.2% (37.3; 72.2) |
| | No | 107 | 46 (43%) | 61 (57%) | 719 (467; 878)[2] | |
| Renal disease | Yes | 297 | 89 (30%) | 208 (70%) | 99 (66; 226) | 44.0% (21.2; 60.2) |
| | No | 203 | 82 (40%) | 121 (60%) | 732 (554; 885)[2] | |
| Type 2 diabetes | Yes | 576 | 137 (24%) | 439 (76%) | 108 (67; 227) | 40.9% (26.8; 52.3) |
| | No | 355 | 123 (35%) | 232 (65%) | 718 (541; 882)[2] | |
| Immunodeficiency or cancer [3] | Yes | 316 | 93 (29%) | 223 (71%) | 144 (60; 233) | 46.6% (-24.6; 77.1) |
| | No | 332 | 125 (38%) | 207 (62%) | 728 (534; 872)[2] | |
| **Number of chronic conditions** | | | | | | |
| 0-1[4] | Yes | 383 | 103 (27%) | 280 (73%) | 134 (68; 228) | 31.8% (18.2; 43.2) |
| | No | 583 | 160 (27%) | 423 (73%) | 775 (648; 926)[2] | |
| 2 | Yes | 488 | 95 (19%) | 393 (81%) | 107 (67; 221) | 69.8% (64.7; 74.1) |
| | No | 339 | 112 (33%) | 227 (67%) | 737 (543; 893)[2] | |
| ≥3 | Yes | 762 | 180 (24%) | 582 (76%) | 100 (63; 220) | 42.1% (31.2; 51.2) |
| | No | 504 | 177 (35%) | 327 (65%) | 710 (489; 834)[2] | |
| **Weeks since BNT162b2 XBB vaccine dose** | | | | | | |
| 2 to <30 wks | Yes | 1146 | 193 (17%) | 953 (83%) | 77 (55; 120) | 64.5% (56.6; 71.0) |
| | No | 1426 | 449 (31%) | 977 (69%) | 744 (566; 898)[2] | |
| 2 to <12 wks | Yes | 660 | 105 (16%) | 555 (84%) | 59 (45; 71) | 68.5% (55.9; 77.6) |
| | No | 1426 | 449 (31%) | 977 (69%) | 744 (566; 898)[2] | |
| 12 to <18 wks | Yes | 212 | 28 (13%) | 184 (87%) | 96 (89; 107) | 75.3% (68.5; 80.6) |
| | No | 1426 | 449 (31%) | 977 (69%) | 744 (566; 898)[2] | |
| 18 to <24 wks | Yes | 101 | 20 (20%) | 81 (80%) | 145 (137; 155) | 59.3% (44.1; 70.4) |
| | No | 1426 | 449 (31%) | 977 (69%) | 744 (566; 898)[2] | |
| 24 to <30 wks | Yes | 173 | 40 (23%) | 133 (77%) | 194 (182; 203) | 50.7% (35.7; 62.2) |
| | No | 1426 | 449 (31%) | 977 (69%) | 744 (566; 898)[2] | |
| 30 to <46 wks | Yes | 487 | 185 (38%) | 302 (62%) | 250 (230; 272) | 4.5% (-31.3; 30.5) |
| | No | 1426 | 449 (31%) | 977 (69%) | 744 (566; 898)[2] | |
| **Severity level[5]** | | | | | | |
| Hospital admission, no ICU, no IH death | Yes | 1489 | 347 (23%) | 1142 (77%) | 110 (66; 223) | 46.5% (31.8; 58.1) |
| | No | 1304 | 408 (31%) | 896 (69%) | 747 (566; 898)[2] | |
| ICU admission, no IH death | Yes | 29 | 4 (14%) | 25 (86%) | 77 (62; 215) | 63.7% (23.8; 82.7)* |
| | No | 50 | 15 (30%) | 35 (70%) | 778 (695; 882)[2] | |
| In-hospital death | Yes | 113 | 26 (23%) | 87 (77%) | 88 (62; 224) | 57.8% (-15.3; 84.5) |
| | No | 68 | 24 (35%) | 44 (65%) | 645 (525; 904)[2] | |
| ICU admission or IH death | Yes | 142 | 30 (21%) | 112 (79%) | 86 (62; 216) | 55.4% (21.3; 74.8) |
| | No | 118 | 39 (33%) | 79 (67%) | 716 (555; 900)[2] | |
| **Respiratory support level[6]** | | | | | | |
| None | Yes | 284 | 69 (24%) | 215 (76%) | 114 (62; 226) | 61.9% (46.3; 73.0) |
| | No | 308 | 98 (32%) | 210 (68%) | 778 (623; 918)[2] | |
| Oxygen therapy | Yes | 901 | 187 (21%) | 714 (79%) | 121 (68; 224) | 50.5% (31.3; 64.3) |
| | No | 596 | 187 (31%) | 409 (69%) | 737 (568; 899)[2] | |
| Non-invasive ventilation | Yes | 174 | 34 (20%) | 140 (80%) | 97 (61; 216) | -38.5% (-88.3; -1.8) |
| | No | 96 | 19 (20%) | 77 (80%) | 724 (479; 901)[2] | |
| Invasive mechanical ventilation | Yes | 9 | 0 (0%) | 9 (100%) | 69 (61; 97) | |
| | No | 10 | 3 (30%) | 7 (70%) | 722 (708; 786)[2] | |
| ECMO | Yes | 0 | 0 (0%) | 0 (0%) | | |
| | No | 1 | 1 (100%) | 0 (0%) | 519 (519; 519)[2] | |
| **Length of stay** | | | | | | |
| LOS ≥ 2 days | Yes | 1603 | 368 (23%) | 1235 (77%) | 107 (65; 223) | 49.2% (40.6; 56.5) |
| | No | 1379 | 436 (32%) | 943 (68%) | 744 (566; 899)[2] | |
| LOS ≥ 4 days | Yes | 1296 | 295 (23%) | 1001 (77%) | 102 (64; 222) | 50.4% (43.3; 56.6) |
| | No | 1113 | 359 (32%) | 754 (68%) | 744 (566; 898)[2] | |
| LOS ≥ 7 days | Yes | 692 | 137 (20%) | 555 (80%) | 88 (61; 210) | 61.5% (51.7; 69.3) |
| | No | 635 | 197 (31%) | 438 (69%) | 735 (563; 875)[2] | |

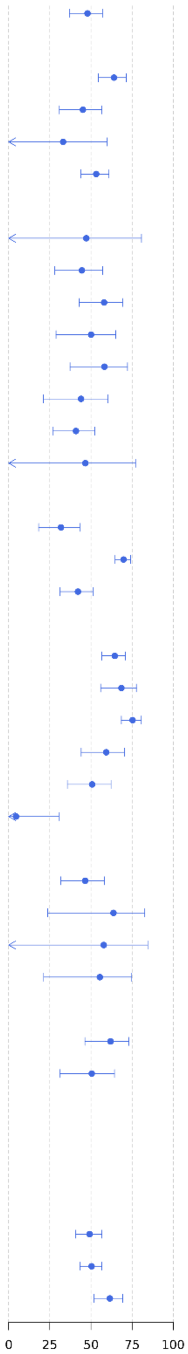

*Abbreviations: ECMO, extracorporeal membrane oxygenation; ICU, intensive care unit; IH, in-hospital; N, number; SARI, severe acute respiratory infections.*

*[1] Vaccine effectiveness (VE) estimates are adjusted for date of symptom onset, age, sex, number of chronic conditions and receipt of influenza vaccine in the 12 months prior to current hospital admission.*

*[2] 'Never vaccinated' subjects were excluded when calculating the median (interquartile range) of time since last vaccine dose in the unexposed group (patients who did not receive any dose of a COVID-19 vaccine in the 2023–2024 autumn/winter season).*

*[3] The categories "Cancer" and "Immunodeficiency" among chronic conditions are counted as separate types of chronic conditions in all adjusted VE estimates. Here, the VE estimate among participants having "Immunodeficiency or cancer" is provided as a composite.*

*[4] The categories "0" and "1" among the number of chronic conditions are counted as separate categories of number of chronic conditions in all adjusted VE estimates. Here, the VE estimate among participants having "0–1"chronic conditions is provided as a composite.*

*[5] The categories "Hospital admission, no ICU, no IH death", "ICU admission, no IH death", and "In-hospital death" are mutually exclusive. An additional category is added to combine the latter two categories: "ICU admission or IH death".*

*[6] Respiratory support level, presented in ascending order of severity with the highest level used during hospital stay determine the category, are mutually exclusive: 1) none;, 2) oxygen therapy (e.g., nasal cannula or mask); 3) non-invasive ventilation (support without endotracheal intubation such as high flow nasal oxygen, continuous or bi-level positive airway pressure); 4) invasive mechanical ventilation (support with endotracheal intubation); and 5) extracorporeal membrane oxygenation.*

#### Sensitivity analysis with regard to location and number of knots of age

**Supplementary Figure S6 [Sensitivity analysis 2a]** Vaccine effectiveness against JN.1-related hospitalisation in SARI patients who received at least one dose of BNT162b2 XBB vaccine compared to patients who did not receive any dose of a COVID-19 vaccine in the 2023–2024 autumn/winter season. The location of knots for ages were chosen at 60 and 75 years.

| Strata | Exposure | Total, N | Test-positive cases: N (%) | Test-negative controls: N (%) | Days since last vaccine dose: Median (Q1; Q3) | VE (95% CI)[1] |
|---|---|---|---|---|---|---|
| **Overall** | | | | | | |
| Overall | Yes | 1633 | 378 (23%) | 1255 (77%) | 107 (65; 223) | 48.5% (38.3; 57.1) |
| | No | 1426 | 449 (31%) | 977 (69%) | 744 (566; 898)[2] | |
| **Age group (years)** | | | | | | |
| 18-64y | Yes | 166 | 35 (21%) | 131 (79%) | 106 (62; 226) | 65.6% (55.5; 73.5) |
| | No | 525 | 153 (29%) | 372 (71%) | 791 (701; 916)[2] | |
| ≥65y | Yes | 1467 | 343 (23%) | 1124 (77%) | 107 (65; 223) | 44.9% (39.5; 49.7) |
| | No | 901 | 296 (33%) | 605 (67%) | 717 (510; 846)[2] | |
| 65-79y | Yes | 589 | 129 (22%) | 460 (78%) | 115 (60; 220) | 33.1% (-11.3; 59.8) |
| | No | 457 | 132 (29%) | 325 (71%) | 724 (533; 873)[2] | |
| ≥80y | Yes | 878 | 214 (24%) | 664 (76%) | 104 (68; 224) | 53.2% (43.9; 60.9) |
| | No | 444 | 164 (37%) | 280 (63%) | 692 (476; 836)[2] | |
| **Type of chronic condition** | | | | | | |
| Asthma | Yes | 139 | 26 (19%) | 113 (81%) | 86 (60; 204) | 45.8% (-44.1; 79.6) |
| | No | 130 | 37 (28%) | 93 (72%) | 737 (556; 898)[2] | |
| Lung disease | Yes | 649 | 125 (19%) | 524 (81%) | 99 (63; 216) | 44.5% (28.1; 57.1) |
| | No | 440 | 128 (29%) | 312 (71%) | 723 (496; 846)[2] | |
| Cardiovascular disease | Yes | 780 | 170 (22%) | 610 (78%) | 95 (63; 220) | 58.5% (43.7; 69.5) |
| | No | 503 | 179 (36%) | 324 (64%) | 723 (507; 845)[2] | |
| Hypertension | Yes | 1161 | 258 (22%) | 903 (78%) | 102 (64; 220) | 50.7% (30.1; 65.3) |
| | No | 747 | 258 (35%) | 489 (65%) | 724 (519; 848)[2] | |
| Chronic liver disease | Yes | 89 | 25 (28%) | 64 (72%) | 131 (66; 224) | 58.2% (35.5; 73.0) |
| | No | 107 | 46 (43%) | 61 (57%) | 719 (467; 878)[2] | |
| Renal disease | Yes | 297 | 89 (30%) | 208 (70%) | 99 (66; 226) | 48.4% (26.6; 63.8) |
| | No | 203 | 82 (40%) | 121 (60%) | 732 (554; 885)[2] | |
| Type 2 diabetes | Yes | 576 | 137 (24%) | 439 (76%) | 108 (67; 227) | 43.4% (28.6; 55.1) |
| | No | 355 | 123 (35%) | 232 (65%) | 718 (541; 882)[2] | |
| Immunodeficiency or cancer [3] | Yes | 316 | 93 (29%) | 223 (71%) | 144 (60; 233) | 46.8% (-23.9; 77.1) |
| | No | 332 | 125 (38%) | 207 (62%) | 728 (534; 872)[2] | |
| **Number of chronic conditions** | | | | | | |
| 0-1[4] | Yes | 383 | 103 (27%) | 280 (73%) | 134 (68; 228) | 32.0% (18.4; 43.3) |
| | No | 583 | 160 (27%) | 423 (73%) | 775 (648; 926)[2] | |
| 2 | Yes | 488 | 95 (19%) | 393 (81%) | 107 (67; 221) | 69.6% (64.5; 74.0) |
| | No | 339 | 112 (33%) | 227 (67%) | 737 (543; 893)[2] | |
| ≥3 | Yes | 762 | 180 (24%) | 582 (76%) | 100 (63; 220) | 44.3% (35.5; 51.9) |
| | No | 504 | 177 (35%) | 327 (65%) | 710 (489; 834)[2] | |
| **Weeks since BNT162b2 XBB vaccine dose** | | | | | | |
| 2 to <30 wks | Yes | 1146 | 193 (17%) | 953 (83%) | 77 (55; 120) | 65.2% (57.5; 71.5) |
| | No | 1426 | 449 (31%) | 977 (69%) | 744 (566; 898)[2] | |
| 2 to <12 wks | Yes | 660 | 105 (16%) | 555 (84%) | 59 (45; 71) | 69.2% (57.1; 77.9) |
| | No | 1426 | 449 (31%) | 977 (69%) | 744 (566; 898)[2] | |
| 12 to <18 wks | Yes | 212 | 28 (13%) | 184 (87%) | 96 (89; 107) | 75.8% (68.9; 81.2) |
| | No | 1426 | 449 (31%) | 977 (69%) | 744 (566; 898)[2] | |
| 18 to <24 wks | Yes | 101 | 20 (20%) | 81 (80%) | 145 (137; 155) | 60.0% (44.1; 71.4) |
| | No | 1426 | 449 (31%) | 977 (69%) | 744 (566; 898)[2] | |
| 24 to <30 wks | Yes | 173 | 40 (23%) | 133 (77%) | 194 (182; 203) | 50.6% (36.4; 61.6) |
| | No | 1426 | 449 (31%) | 977 (69%) | 744 (566; 898)[2] | |
| 30 to <46 wks | Yes | 487 | 185 (38%) | 302 (62%) | 250 (230; 272) | 5.7% (-29.3; 31.3) |
| | No | 1426 | 449 (31%) | 977 (69%) | 744 (566; 898)[2] | |
| **Severity level[5]** | | | | | | |
| Hospital admission, no ICU, no IH death | Yes | 1489 | 347 (23%) | 1142 (77%) | 110 (66; 223) | 47.1% (33.1; 58.2) |
| | No | 1304 | 408 (31%) | 896 (69%) | 747 (566; 898)[2] | |
| ICU admission, no IH death | Yes | 29 | 4 (14%) | 25 (86%) | 77 (62; 215) | 65.0% (20.8; 84.6) |
| | No | 50 | 15 (30%) | 35 (70%) | 778 (695; 882)[2] | |
| In-hospital death | Yes | 113 | 26 (23%) | 87 (77%) | 88 (62; 224) | 59.1% (-19.1; 86.0) |
| | No | 68 | 24 (35%) | 44 (65%) | 645 (525; 904)[2] | |
| ICU admission or IH death | Yes | 142 | 30 (21%) | 112 (79%) | 86 (62; 216) | 58.4% (18.2; 78.8) |
| | No | 118 | 39 (33%) | 79 (67%) | 716 (555; 900)[2] | |
| **Respiratory support level[6]** | | | | | | |
| None | Yes | 284 | 69 (24%) | 215 (76%) | 114 (62; 226) | 61.7% (46.3; 72.7) |
| | No | 308 | 98 (32%) | 210 (68%) | 778 (623; 918)[2] | |
| Oxygen therapy | Yes | 901 | 187 (21%) | 714 (79%) | 121 (68; 224) | 51.0% (31.9; 64.8) |
| | No | 596 | 187 (31%) | 409 (69%) | 737 (568; 899)[2] | |
| Non-invasive ventilation | Yes | 174 | 34 (20%) | 140 (80%) | 97 (61; 216) | -31.0% (-73.4; 1.1) |
| | No | 96 | 19 (20%) | 77 (80%) | 724 (479; 901)[2] | |
| Invasive mechanical ventilation | Yes | 9 | 0 (0%) | 9 (100%) | 69 (61; 97) | |
| | No | 10 | 3 (30%) | 7 (70%) | 722 (708; 786)[2] | |
| ECMO | Yes | 0 | 0 (0%) | 0 (0%) | | |
| | No | 1 | 1 (100%) | 0 (0%) | 519 (519; 519)[2] | |
| **Length of stay** | | | | | | |
| LOS ≥ 2 days | Yes | 1603 | 368 (23%) | 1235 (77%) | 107 (65; 223) | 49.9% (41.8; 56.8) |
| | No | 1379 | 436 (32%) | 943 (68%) | 744 (566; 899)[2] | |
| LOS ≥ 4 days | Yes | 1296 | 295 (23%) | 1001 (77%) | 102 (64; 222) | 51.1% (43.7; 57.6) |
| | No | 1113 | 359 (32%) | 754 (68%) | 744 (566; 898)[2] | |
| LOS ≥ 7 days | Yes | 692 | 137 (20%) | 555 (80%) | 88 (61; 210) | 62.7% (52.5; 70.8) |
| | No | 635 | 197 (31%) | 438 (69%) | 735 (563; 875)[2] | |

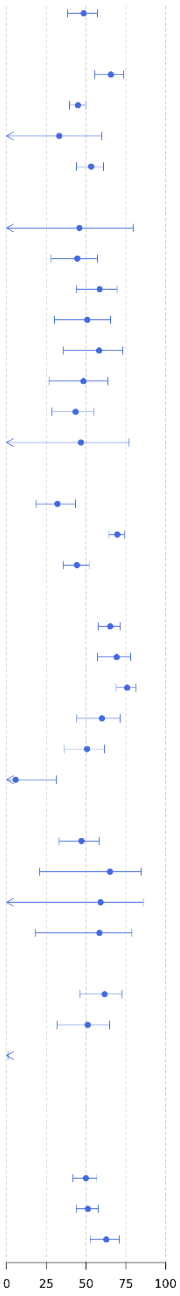

*Abbreviations: ECMO, extracorporeal membrane oxygenation; ICU, intensive care unit; IH, in-hospital; N, number; SARI, severe acute respiratory infections.*

[1] *Vaccine effectiveness (VE) estimates are adjusted for date of symptom onset, age, sex, number of chronic conditions and receipt of influenza vaccine in the 12 months prior to current hospital admission.*

[2] *'Never vaccinated' subjects were excluded when calculating the median (interquartile range) of time since last vaccine dose in the unexposed group (patients who did not receive any dose of a COVID-19 vaccine in the 2023–2024 autumn/winter season).*

[3] *The categories "Cancer" and "Immunodeficiency" among chronic conditions are counted as separate types of chronic conditions in all adjusted VE estimates. Here, the VE estimate among participants having "Immunodeficiency or cancer" is provided as a composite.*

[4] *The categories "0" and "1" among the number of chronic conditions are counted as separate categories of number of chronic conditions in all adjusted VE estimates. Here, the VE estimate among participants having "0–1"chronic conditions is provided as a composite.*

[5] *The categories "Hospital admission, no ICU, no IH death", "ICU admission, no IH death", and "In-hospital death" are mutually exclusive. An additional category is added to combine the latter two categories: "ICU admission or IH death".*

[6] *Respiratory support level, presented in ascending order of severity with the highest level used during hospital stay determine the category, are mutually exclusive: 1) none;, 2) oxygen therapy (e.g., nasal cannula or mask); 3) non-invasive ventilation (support without endotracheal intubation such as high flow nasal oxygen, continuous or bi-level positive airway pressure); 4) invasive mechanical ventilation (support with endotracheal intubation); and 5) extracorporeal membrane oxygenation.*

**Supplementary Figure S7 [Sensitivity analysis 2b]** Vaccine effectiveness against JN.1-related hospitalisation in SARI patients who received at least one dose of BNT162b2 XBB vaccine compared to patients who did not receive any dose of a COVID-19 vaccine in the 2023–2024 autumn/winter season. The location of knots for ages were chosen at 50, 65, and 80 years.

| Strata | Exposure | Total, N | Test-positive cases: N (%) | Test-negative controls: N (%) | Days since last vaccine dose: Median (Q1; Q3) | VE (95% CI)[1] |
|---|---|---|---|---|---|---|
| **Overall** | | | | | | |
| Overall | Yes | 1633 | 378 (23%) | 1255 (77%) | 107 (65; 223) | 48.5% (38.2; 57.0) |
| | No | 1426 | 449 (31%) | 977 (69%) | 744 (566; 898)[2] | |
| **Age group (years)** | | | | | | |
| 18-64y | Yes | 166 | 35 (21%) | 131 (79%) | 106 (62; 226) | 65.6% (55.5; 73.5) |
| | No | 525 | 153 (29%) | 372 (71%) | 791 (701; 916)[2] | |
| ≥65y | Yes | 1467 | 343 (23%) | 1124 (77%) | 107 (65; 223) | 44.9% (39.5; 49.7) |
| | No | 901 | 296 (33%) | 605 (67%) | 717 (510; 846)[7] | |
| 65-79y | Yes | 589 | 129 (22%) | 460 (78%) | 115 (60; 220) | 33.1% (-11.3; 59.8) |
| | No | 457 | 132 (29%) | 325 (71%) | 724 (533; 873)[7] | |
| ≥80y | Yes | 878 | 214 (24%) | 664 (76%) | 104 (68; 224) | 53.2% (43.9; 60.9) |
| | No | 444 | 164 (37%) | 280 (63%) | 692 (476; 836)[2] | |
| **Type of chronic condition** | | | | | | |
| Asthma | Yes | 139 | 26 (19%) | 113 (81%) | 86 (60; 204) | 44.7% (-56.2; 80.4) |
| | No | 130 | 37 (28%) | 93 (72%) | 737 (556; 898)[2] | |
| Lung disease | Yes | 649 | 125 (19%) | 524 (81%) | 99 (63; 216) | 44.5% (28.0; 57.2) |
| | No | 440 | 128 (29%) | 312 (71%) | 723 (496; 846)[4] | |
| Cardiovascular disease | Yes | 780 | 170 (22%) | 610 (78%) | 95 (63; 220) | 58.6% (43.9; 69.5) |
| | No | 503 | 179 (36%) | 324 (64%) | 723 (507; 845)[4] | |
| Hypertension | Yes | 1161 | 258 (22%) | 903 (78%) | 102 (64; 220) | 50.8% (30.4; 65.2) |
| | No | 747 | 258 (35%) | 489 (65%) | 724 (519; 848)[2] | |
| Chronic liver disease | Yes | 89 | 25 (28%) | 64 (72%) | 131 (66; 224) | 58.9% (37.4; 73.0) |
| | No | 107 | 46 (43%) | 61 (57%) | 719 (467; 878)[2] | |
| Renal disease | Yes | 297 | 89 (30%) | 208 (70%) | 99 (66; 226) | 48.2% (24.5; 64.5) |
| | No | 203 | 82 (40%) | 121 (60%) | 732 (554; 885)[7] | |
| Type 2 diabetes | Yes | 576 | 137 (24%) | 439 (76%) | 108 (67; 227) | 43.1% (28.8; 54.5) |
| | No | 355 | 123 (35%) | 232 (65%) | 718 (541; 882)[7] | |
| Immunodeficiency or cancer [3] | Yes | 316 | 93 (29%) | 223 (71%) | 144 (60; 233) | 46.6% (-24.9; 77.2) |
| | No | 332 | 125 (38%) | 207 (62%) | 728 (534; 872)[7] | |
| **Number of chronic conditions** | | | | | | |
| 0-1[4] | Yes | 383 | 103 (27%) | 280 (73%) | 134 (68; 228) | 32.3% (18.6; 43.7) |
| | No | 583 | 160 (27%) | 423 (73%) | 775 (648; 926)[2] | |
| 2 | Yes | 488 | 95 (19%) | 393 (81%) | 107 (67; 221) | 70.0% (64.7; 74.5) |
| | No | 339 | 112 (33%) | 227 (67%) | 737 (543; 893)[2] | |
| ≥3 | Yes | 762 | 180 (24%) | 582 (76%) | 100 (63; 220) | 44.4% (35.0; 52.5) |
| | No | 504 | 177 (35%) | 327 (65%) | 710 (489; 834)[2] | |
| **Weeks since BNT162b2 XBB vaccine dose** | | | | | | |
| 2 to <30 wks | Yes | 1146 | 193 (17%) | 953 (83%) | 77 (55; 120) | 65.0% (57.3; 71.4) |
| | No | 1426 | 449 (31%) | 977 (69%) | 744 (566; 898)[2] | |
| 2 to <12 wks | Yes | 660 | 105 (16%) | 555 (84%) | 59 (45; 71) | 69.0% (57.0; 77.6) |
| | No | 1426 | 449 (31%) | 977 (69%) | 744 (566; 898)[2] | |
| 12 to <18 wks | Yes | 212 | 28 (13%) | 184 (87%) | 96 (89; 107) | 75.7% (68.6; 81.2) |
| | No | 1426 | 449 (31%) | 977 (69%) | 744 (566; 898)[7] | |
| 18 to <24 wks | Yes | 101 | 20 (20%) | 81 (80%) | 145 (137; 155) | 60.0% (44.0; 71.4) |
| | No | 1426 | 449 (31%) | 977 (69%) | 744 (566; 898)[7] | |
| 24 to <30 wks | Yes | 173 | 40 (23%) | 133 (77%) | 194 (182; 203) | 50.6% (36.6; 61.6) |
| | No | 1426 | 449 (31%) | 977 (69%) | 744 (566; 898)[2] | |
| 30 to <46 wks | Yes | 487 | 185 (38%) | 302 (62%) | 250 (230; 272) | 6.0% (-28.6; 31.2) |
| | No | 1426 | 449 (31%) | 977 (69%) | 744 (566; 898)[2] | |
| **Severity level[5]** | | | | | | |
| Hospital admission, no ICU, no IH death | Yes | 1489 | 347 (23%) | 1142 (77%) | 110 (66; 223) | 47.1% (33.0; 58.1) |
| | No | 1304 | 408 (31%) | 896 (69%) | 747 (566; 898)[2] | |
| ICU admission, no IH death | Yes | 29 | 4 (14%) | 25 (86%) | 77 (62; 215) | NE* |
| | No | 50 | 15 (30%) | 35 (70%) | 778 (695; 882)[2] | |
| In-hospital death | Yes | 113 | 26 (23%) | 87 (77%) | 88 (62; 224) | 58.9% (-15.0; 85.3) |
| | No | 68 | 24 (35%) | 44 (65%) | 645 (525; 904)[4] | |
| ICU admission or IH death | Yes | 142 | 30 (21%) | 112 (79%) | 86 (62; 216) | 59.8% (16.8; 80.6) |
| | No | 118 | 39 (33%) | 79 (67%) | 716 (555; 900)[4] | |
| **Respiratory support level[6]** | | | | | | |
| None | Yes | 284 | 69 (24%) | 215 (76%) | 114 (62; 226) | 61.1% (46.3; 71.8) |
| | No | 308 | 98 (32%) | 210 (68%) | 778 (623; 918)[2] | |
| Oxygen therapy | Yes | 901 | 187 (21%) | 714 (79%) | 121 (68; 224) | 51.0% (32.0; 64.7) |
| | No | 596 | 187 (31%) | 409 (69%) | 737 (568; 899)[2] | |
| Non-invasive ventilation | Yes | 174 | 34 (20%) | 140 (80%) | 97 (61; 216) | -34.2% (-79.1; -0.5) |
| | No | 96 | 19 (20%) | 77 (80%) | 724 (479; 901)[7] | |
| Invasive mechanical ventilation | Yes | 9 | 0 (0%) | 9 (100%) | 69 (61; 97) | |
| | No | 10 | 3 (30%) | 7 (70%) | 722 (708; 786)[7] | |
| ECMO | Yes | 0 | 0 (0%) | 0 (0%) | | |
| | No | 1 | 1 (100%) | 0 (0%) | 519 (519; 519)[2] | |
| **Length of stay** | | | | | | |
| LOS ≥ 2 days | Yes | 1603 | 368 (23%) | 1235 (77%) | 107 (65; 223) | 49.8% (41.7; 56.8) |
| | No | 1379 | 436 (32%) | 943 (68%) | 744 (566; 899)[4] | |
| LOS ≥ 4 days | Yes | 1296 | 295 (23%) | 1001 (77%) | 102 (64; 222) | 51.1% (43.6; 57.6) |
| | No | 1113 | 359 (32%) | 754 (68%) | 744 (566; 898)[4] | |
| LOS ≥ 7 days | Yes | 692 | 137 (20%) | 555 (80%) | 88 (61; 210) | 62.8% (52.5; 70.9) |
| | No | 635 | 197 (31%) | 438 (69%) | 735 (563; 875)[4] | |

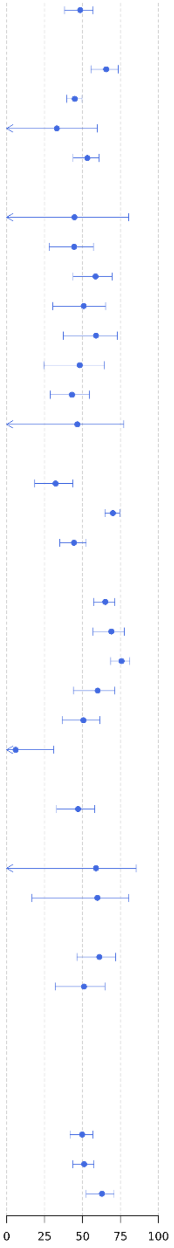

*Abbreviations: ECMO, extracorporeal membrane oxygenation; ICU, intensive care unit; IH, in-hospital; N, number; SARI, severe acute respiratory infections.*

[1] *Vaccine effectiveness (VE) estimates are adjusted for date of symptom onset, age, sex, number of chronic conditions and receipt of influenza vaccine in the 12 months prior to current hospital admission.*

[2] *'Never vaccinated' subjects were excluded when calculating the median (interquartile range) of time since last vaccine dose in the unexposed group (patients who did not receive any dose of a COVID-19 vaccine in the 2023–2024 autumn/winter season).*

[3] *The categories "Cancer" and "Immunodeficiency" among chronic conditions are counted as separate types of chronic conditions in all adjusted VE estimates. Here, the VE estimate among participants having "Immunodeficiency or cancer" is provided as a composite.*

[4] *The categories "0" and "1" among the number of chronic conditions are counted as separate categories of number of chronic conditions in all adjusted VE estimates. Here, the VE estimate among participants having "0–1"chronic conditions is provided as a composite.*

[5] *The categories "Hospital admission, no ICU, no IH death", "ICU admission, no IH death", and "In-hospital death" are mutually exclusive. An additional category is added to combine the latter two categories: "ICU admission or IH death".*

[6] *Respiratory support level, presented in ascending order of severity with the highest level used during hospital stay determine the category, are mutually exclusive: 1) none;, 2) oxygen therapy (e.g., nasal cannula or mask); 3) non-invasive ventilation (support without endotracheal intubation such as high flow nasal oxygen, continuous or bi-level positive airway pressure); 4) invasive mechanical ventilation (support with endotracheal intubation); and 5) extracorporeal membrane oxygenation.*

** Due to limited number of subjects the VE estimates have a wide 95% CI leading to unreliable interpretation of the results, hence results are not shown*

#### Sensitivity analysis with regard to VE estimates stratified by history of prior COVID-19 infection

**Supplementary Figure S8 [Sensitivity analysis 3]** Vaccine effectiveness against JN.1-related hospitalisation in SARI patients who received at least one dose of BNT162b2 XBB vaccine compared to patients who did not receive any dose of a COVID-19 vaccine in the 2023–2024 autumn/winter season. Results for VE estimates stratified by history of prior COVID-19 infection are presented.

| Strata | Exposure | Total, N | Test-positive cases: N (%) | Test-negative controls: N (%) | Days since last vaccine dose: Median (Q1; Q3) | VE (95% CI)[1] |
|---|---|---|---|---|---|---|
| **Overall** | | | | | | |
| Overall | Yes | 1633 | 378 (23%) | 1255 (77%) | 107 (65; 223) | 47.9% (37.0; 56.9) |
| | No | 1426 | 449 (31%) | 977 (69%) | 744 (566; 898)[2] | |
| **Prior COVID-19 history** | | | | | | |
| Prior COVID-19 infection | Yes | 520 | 124 (24%) | 396 (76%) | 180 (76; 239) | 39.8% (12.4; 58.6) |
| | No | 393 | 119 (30%) | 274 (70%) | 769 (604; 926)[2] | |
| No prior COVID-19 infection | Yes | 940 | 218 (23%) | 722 (77%) | 112 (65; 225) | 46.4% (36.0; 55.1) |
| | No | 777 | 257 (33%) | 520 (67%) | 755 (574; 910)[2] | |

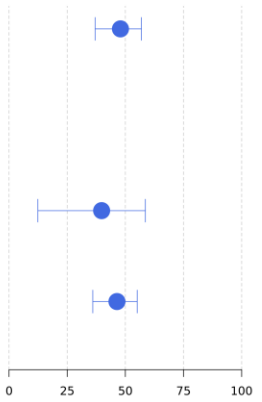

*Abbreviations: N, number; SARI, severe acute respiratory infections.*

[1] *Vaccine effectiveness (VE) estimates are adjusted for date of symptom onset, age, sex, number of chronic conditions and receipt of influenza vaccine in the 12 months prior to current hospital admission.*

[2] *'Never vaccinated' subjects were excluded when calculating the median (interquartile range) of time since last vaccine dose in the unexposed group (patients who did not receive any dose of a COVID-19 vaccine in the 2023–2024 autumn/winter season).*

#### Sensitivity analysis with regard to VE estimates while not accounting for influenza vaccination status

**Supplementary Figure S9 [Sensitivity analysis 4]** Vaccine effectiveness against JN.1-related hospitalisation in SARI patients who received at least one dose of BNT162b2 XBB vaccine compared to patients who did not receive any dose of a COVID-19 vaccine in the 2023–2024 autumn/winter, adjusted only for symptom onset date, age, sex, and number of chronic conditions.

| Strata | Exposure | Total, N | Test-positive cases: N (%) | Test-negative controls: N (%) | Days since last vaccine dose: Median (Q1; Q3) | VE (95% CI)[1] |
|---|---|---|---|---|---|---|
| **Overall** | | | | | | |
| Overall | Yes | 1633 | 378 (23%) | 1255 (77%) | 107 (65; 223) | 40.9% (24.3; 53.9) |
| | No | 1426 | 449 (31%) | 977 (69%) | 744 (566; 898)[2] | |
| **Age group (years)** | | | | | | |
| 18-64y | Yes | 166 | 35 (21%) | 131 (79%) | 106 (62; 226) | 51.8% (39.3; 61.7) |
| | No | 525 | 153 (29%) | 372 (71%) | 791 (701; 916)[2] | |
| ≥65y | Yes | 1467 | 343 (23%) | 1124 (77%) | 107 (65; 223) | 38.2% (25.6; 48.6) |
| | No | 901 | 296 (33%) | 605 (67%) | 717 (510; 846)[2] | |
| 65-79y | Yes | 589 | 129 (22%) | 460 (78%) | 115 (60; 220) | 30.6% (0.9; 51.4) |
| | No | 457 | 132 (29%) | 325 (71%) | 724 (533; 873)[2] | |
| ≥80y | Yes | 878 | 214 (24%) | 664 (76%) | 104 (68; 224) | 45.9% (33.3; 56.1) |
| | No | 444 | 164 (37%) | 280 (63%) | 692 (476; 836)[2] | |
| **Type of chronic condition** | | | | | | |
| Asthma | Yes | 139 | 26 (19%) | 113 (81%) | 86 (60; 204) | 37.4% (21.8; 49.9) |
| | No | 130 | 37 (28%) | 93 (72%) | 737 (556; 898)[2] | |
| Lung disease | Yes | 649 | 125 (19%) | 524 (81%) | 99 (63; 216) | 43.4% (34.6; 51.1) |
| | No | 440 | 128 (29%) | 312 (71%) | 723 (496; 846)[2] | |
| Cardiovascular disease | Yes | 780 | 170 (22%) | 610 (78%) | 95 (63; 220) | 52.3% (31.6; 66.7) |
| | No | 503 | 179 (36%) | 324 (64%) | 723 (507; 845)[2] | |
| Hypertension | Yes | 1161 | 258 (22%) | 903 (78%) | 102 (64; 220) | 47.8% (25.8; 63.3) |
| | No | 747 | 258 (35%) | 489 (65%) | 724 (519; 848)[2] | |
| Chronic liver disease | Yes | 89 | 25 (28%) | 64 (72%) | 131 (66; 224) | 47.4% (7.4; 70.1) |
| | No | 107 | 46 (43%) | 61 (57%) | 719 (467; 878)[2] | |
| Renal disease | Yes | 297 | 89 (30%) | 208 (70%) | 99 (66; 226) | 22.3% (7.2; 34.9) |
| | No | 203 | 82 (40%) | 121 (60%) | 732 (554; 885)[2] | |
| Type 2 diabetes | Yes | 576 | 137 (24%) | 439 (76%) | 108 (67; 227) | 42.2% (30.4; 52.0) |
| | No | 355 | 123 (35%) | 232 (65%) | 718 (541; 882)[2] | |
| Immunodeficiency or cancer [3] | Yes | 316 | 93 (29%) | 223 (71%) | 144 (60; 233) | 27.6% (-10.6; 52.5) |
| | No | 332 | 125 (38%) | 207 (62%) | 728 (534; 872)[2] | |
| **Number of chronic conditions** | | | | | | |
| 0-1[4] | Yes | 383 | 103 (27%) | 280 (73%) | 134 (68; 228) | 18.5% (-0.8; 34.1) |
| | No | 583 | 160 (27%) | 423 (73%) | 775 (648; 926)[2] | |
| 2 | Yes | 488 | 95 (19%) | 393 (81%) | 107 (67; 221) | 55.3% (32.2; 70.5) |
| | No | 339 | 112 (33%) | 227 (67%) | 737 (543; 893)[2] | |
| ≥3 | Yes | 762 | 180 (24%) | 582 (76%) | 100 (63; 220) | 41.3% (31.6; 49.7) |
| | No | 504 | 177 (35%) | 327 (65%) | 710 (489; 834)[2] | |
| **Weeks since BNT162b2 XBB vaccine dose** | | | | | | |
| 2 to <30 wks | Yes | 1146 | 193 (17%) | 953 (83%) | 77 (55; 120) | 59.5% (46.8; 69.1) |
| | No | 1426 | 449 (31%) | 977 (69%) | 744 (566; 898)[2] | |
| 2 to <12 wks | Yes | 660 | 105 (16%) | 555 (84%) | 59 (45; 71) | 62.3% (44.3; 74.5) |
| | No | 1426 | 449 (31%) | 977 (69%) | 744 (566; 898)[2] | |
| 12 to <18 wks | Yes | 212 | 28 (13%) | 184 (87%) | 96 (89; 107) | 71.4% (70.0; 72.7) |
| | No | 1426 | 449 (31%) | 977 (69%) | 744 (566; 898)[2] | |
| 18 to <24 wks | Yes | 101 | 20 (20%) | 81 (80%) | 145 (137; 155) | 51.9% (36.3; 63.7) |
| | No | 1426 | 449 (31%) | 977 (69%) | 744 (566; 898)[2] | |
| 24 to <30 wks | Yes | 173 | 40 (23%) | 133 (77%) | 194 (182; 203) | 41.1% (6.2; 63.0) |
| | No | 1426 | 449 (31%) | 977 (69%) | 744 (566; 898)[2] | |
| 30 to <46 wks | Yes | 487 | 185 (38%) | 302 (62%) | 250 (230; 272) | -12.3% (-69.3; 25.5) |
| | No | 1426 | 449 (31%) | 977 (69%) | 744 (566; 898)[2] | |
| **Severity level[5]** | | | | | | |
| Hospital admission, no ICU, no IH death | Yes | 1489 | 347 (23%) | 1142 (77%) | 110 (66; 223) | 38.8% (16.0; 55.4) |
| | No | 1304 | 408 (31%) | 896 (69%) | 747 (566; 898)[2] | |
| ICU admission, no IH death | Yes | 29 | 4 (14%) | 25 (86%) | 77 (62; 215) | 79.8% (18.8; 95.0)* |
| | No | 50 | 15 (30%) | 35 (70%) | 778 (695; 882)[2] | |
| In-hospital death | Yes | 113 | 26 (23%) | 87 (77%) | 88 (62; 224) | 57.1% (47.3; 65.0) |
| | No | 68 | 24 (35%) | 44 (65%) | 645 (525; 904)[2] | |
| ICU admission or IH death | Yes | 142 | 30 (21%) | 112 (79%) | 86 (62; 216) | 58.7% (44.4; 69.3) |
| | No | 118 | 39 (33%) | 79 (67%) | 716 (555; 900)[2] | |
| **Respiratory support level[6]** | | | | | | |
| None | Yes | 284 | 69 (24%) | 215 (76%) | 114 (62; 226) | 54.5% (49.8; 58.7) |
| | No | 308 | 98 (32%) | 210 (68%) | 778 (623; 918)[2] | |
| Oxygen therapy | Yes | 901 | 187 (21%) | 714 (79%) | 121 (68; 224) | 47.0% (34.5; 57.2) |
| | No | 596 | 187 (31%) | 409 (69%) | 737 (568; 899)[2] | |
| Non-invasive ventilation | Yes | 174 | 34 (20%) | 140 (80%) | 97 (61; 216) | 2.8% (-8.3; 12.7) |
| | No | 96 | 19 (20%) | 77 (80%) | 724 (479; 901)[2] | |
| Invasive mechanical ventilation | Yes | 9 | 0 (0%) | 9 (100%) | 69 (61; 97) | |
| | No | 10 | 3 (30%) | 7 (70%) | 722 (708; 786)[2] | |
| ECMO | Yes | 0 | 0 (0%) | 0 (0%) | | |
| | No | 1 | 1 (100%) | 0 (0%) | 519 (519; 519)[2] | |
| **Length of stay** | | | | | | |
| LOS ≥ 2 days | Yes | 1603 | 368 (23%) | 1235 (77%) | 107 (65; 223) | 42.3% (28.4; 53.5) |
| | No | 1379 | 436 (32%) | 943 (68%) | 744 (566; 899)[2] | |
| LOS ≥ 4 days | Yes | 1296 | 295 (23%) | 1001 (77%) | 102 (64; 222) | 44.8% (33.6; 54.1) |
| | No | 1113 | 359 (32%) | 754 (68%) | 744 (566; 898)[2] | |
| LOS ≥ 7 days | Yes | 692 | 137 (20%) | 555 (80%) | 88 (61; 210) | 51.6% (43.1; 58.8) |
| | No | 635 | 197 (31%) | 438 (69%) | 735 (563; 875)[2] | |

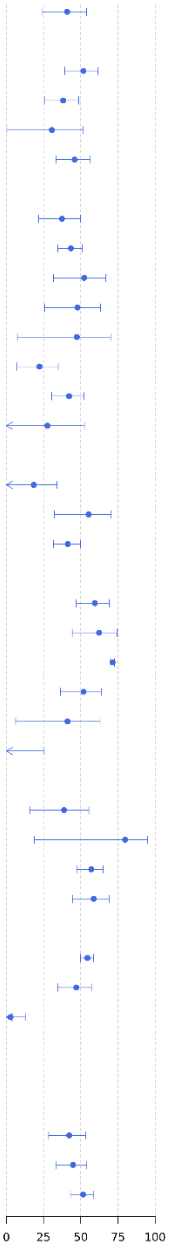

*Abbreviations: ECMO, extracorporeal membrane oxygenation; ICU, intensive care unit; IH, in-hospital; N, number; SARI, severe acute respiratory infections.*

*[1] Vaccine effectiveness (VE) estimates are adjusted for date of symptom onset, age, sex and number of chronic conditions.*

*[2] 'Never vaccinated' subjects were excluded when calculating the median (interquartile range) of time since last vaccine dose in the unexposed group (patients who did not receive any dose of a COVID-19 vaccine in the 2023–2024 autumn/winter season).*

*[3] The categories "Cancer" and "Immunodeficiency" among chronic conditions are counted as separate types of chronic conditions in all adjusted VE estimates. Here, the VE estimate among participants having "Immunodeficiency or cancer" is provided as a composite.*

*[4] The categories "0" and "1" among the number of chronic conditions are counted as separate categories of number of chronic conditions in all adjusted VE estimates. Here, the VE estimate among participants having "0–1"chronic conditions is provided as a composite.*

*[5] The categories "Hospital admission, no ICU, no IH death", "ICU admission, no IH death", and "In-hospital death" are mutually exclusive. An additional category is added to combine the latter two categories: "ICU admission or IH death".*

*[6] Respiratory support level, presented in ascending order of severity with the highest level used during hospital stay determine the category, are mutually exclusive: 1) none;, 2) oxygen therapy (e.g., nasal cannula or mask); 3) non-invasive ventilation (support without endotracheal intubation such as high flow nasal oxygen, continuous or bi-level positive airway pressure); 4) invasive mechanical ventilation (support with endotracheal intubation); and 5) extracorporeal membrane oxygenation.*
